# A novel age-informed approach for genetic association analysis in Alzheimer’s disease

**DOI:** 10.1101/2021.01.05.21249292

**Authors:** Yann Le Guen, Michael E. Belloy, Valerio Napolioni, Sarah J. Eger, Gabriel Kennedy, Ran Tao, Zihuai He, Michael D. Greicius, for the Alzheimer’s Disease Neuroimaging Initiative

## Abstract

**Introduction:** Many Alzheimer’s disease (AD) genetic association studies disregard age or incorrectly account for it, hampering variant discovery.

**Method:** Using simulated data, we compared the statistical power of several models: logistic regression on AD diagnosis adjusted and not adjusted for age; linear regression on a score integrating case-control status and age; and multivariate Cox regression on age-at-onset. We applied these models to real exome-wide data of 11,127 sequenced individuals (54% cases) and replicated suggestive associations in 21,631 genotype-imputed individuals (51% cases).

**Results:** Modelling variable AD risk across age results in 10-20% statistical power gain compared to logistic regression without age adjustment, while incorrect age adjustment leads to critical power loss. Applying our novel AD-age score and/or Cox regression, we discovered and replicated novel variants associated with AD on *KIF21B, USH2A, RAB10, RIN3* and *TAOK2* genes.

**Discussion:** Our AD-age score provides a simple means for statistical power gain and is recommended for future AD studies.

## BACKGROUND

Genetics plays an important role in the onset of Alzheimer’s disease (AD) with an estimated heritability ranging from 58% to 79% [1]. Over the last decade, genome-wide association studies (GWAs) of AD have identified over 40 susceptibility loci [2–5], by meta-analyzing genotype-imputed data from numerous cohorts genotyped on various single nucleotide polymorphism (SNP) arrays. With each updated GWA, the increasing sample sizes and improved imputation quality of low frequency variants have enabled additional discoveries. A complementary approach is to use next generation sequencing to directly genotype every variant, alleviating the need for imputation and enabling rare variant discoveries. To this aim, the Alzheimer’s Disease Sequencing Project (ADSP) undertook whole-exome sequencing (WES) of 10,836 individuals (53% cases) which led to the discovery of novel AD risk genes [6,7]. The ADSP individuals were part of existing AD cohorts and were selected based on a risk score accounting for *APOE* ε2 and *APOE* ε4 alleles, sex, and age-at-onset (AAO) for cases and age at last exam or death for controls [6]. This design promoted the inclusion of controls least likely to develop AD by age 85 years and was shown to maximize statistical power compared to other approaches such as using age matched cases/controls [6].

Across prior AD GWAs, the common approach to association testing was to perform case-control logistic regression analyses adjusted for age. Theoretically, this adjustment should account for increasing AD prevalence with age in the population, independently of genetic factors [8,9]. However, most AD cohorts include the AAO for cases and last known age without cognitive impairment for controls. This common design leads to the average age of cases being lower than the average age of controls. If one performs a case-control logistic regression with a traditional age adjustment, the model will infer that age has a negative effect on AD risk, meaning that younger individuals are more likely to develop AD. Since advanced age is the greatest risk factor for AD [9] it appears essential to correctly account for age. The latter conundrum is particularly relevant to the ADSP where, by design, the average age of controls is 10 years greater than that of cases.

In this work, we aimed to improve on prior AD GWA studies by evaluating and implementing models that inherently, correctly account for age effects on AD. To this aim, we estimated the statistical power of different models on simulated data, reflecting various age differences between cases and controls as found in AD cohorts. These models included logistic regression on AD case-control status adjusted and not adjusted for age, linear regression on a newly designed score which weights case-control status by age, and multivariate Cox regression on AAO, which models cumulative conversion risk across the life span. We then applied these models to exome-wide AD data with a next generation sequenced discovery sample (5,075 controls and 6,052 cases) and replicated suggestive associations in an independent sample of genotype-imputed individuals (10,539 controls and 11,092 cases).

## METHODS

### Power simulations

We performed power simulation studies to evaluate the performance of different AD genetic association models. We first simulated population level data that mimics population AD prevalence estimates at ages 60-100 across a range of age-related risk effect estimates (OR 1.01-1.25) [10,11]. The age effect estimate on AD status (OR 1.16) served as a reference to evaluate power for AD GWA studies [12]. We then simulated AD case-control datasets by random sampling of cases and controls from the population level data. To simulate realistic AD case-control datasets [13–15], subjects’ mean age was centered on 75 years following a binomial distribution with a standard deviation of 8 years. Simulated subjects were restricted to the age range of 60-100, after which cases and controls were randomly drawn abiding by model conditions. To evaluate how age differences between cases and controls affect power for variant discovery, subjects were further sampled to three conditions: 1) no mean age difference between cases and controls, 2) cases’ mean age is 5 years younger than in controls, 3) cases’ mean age is 10 years younger than in controls. These conditions, particularly condition 2, are similar to those observed for common AD GWAS cohorts [13–15], while condition 3 mimics the design of the ADSP WES study. The power was calculated based on 1000 simulation replicates, and the linear regression on the AD-age score was estimated with bootstrap based inference (100 resamplings). Each replicate included either 1000 cases and 1000 controls, or, 5000 cases and 5000 controls, respectively testing for a significance level of α = 0.05, or α = 5×10^−7^ (i.e. exome-wide significance). These parameters respectively mimic common AD GWA cohorts and the ADSP WES study [16]. We evaluated power for a range of realistic effect sizes (OR 1.05, 1.10, 1.20, 1.50) and common minor allele frequency (MAF) 0.01, 0.05-0.45 (at 0.05 increments).

### Participants

All samples were available from publicly released AD-related cohorts, with phenotype and genotype ascertainment described elsewhere [3,6,17–27,13]. The current study protocol was granted an exemption by the Stanford University institutional review board because the analyses were carried out on deidentified, off-the-shelf data; therefore, further informed consent was not required.

The European individuals in ADSP WES [6,7], ADSP whole-genome sequencing (WGS) [22,26] and the Accelerating Medicine Partnership in AD (AMP-AD) WGS [23,25,27] cohorts comprise our discovery sample, and were mega-analyzed (**Tables 1, S1**). The ADSP WES selection criteria have already been introduced, the selection scheme led to a 10 years average age difference between cases and controls [6,16]. For AMP-AD, the reported age for cases was not always AAO; thus, the average age of controls was only two years greater than that of cases.

**Table 1.** Detailed demographics for discovery and replication sample. Details per cohort included in the discovery and replication can be found respectively in **Tables S1** and **S2**. HC: Healthy Controls, AD: Alzheimer’s Disease.

| Sample | *N*<br>(% females) | *Age*<br>$\mu$ ($\sigma$) | $\epsilon_3/\epsilon_3$<br>(%) | $\epsilon_3/\epsilon_4$<br>(%) | $\epsilon_4/\epsilon_4$<br>(%) | $\epsilon_2/\epsilon_3$<br>(%) | $\epsilon_2/\epsilon_4$<br>(%) | $\epsilon_2/\epsilon_2$<br>(%) |
| --- | --- | --- | --- | --- | --- | --- | --- | --- |
| **Discovery – (WES+WGS)** |  |  |  |  |  |  |  |  |
| Controls | 5075 (59.0) | 85.2 (5.4) | 66.13 | 13.93 | 0.51 | 17.12 | 1.52 | 0.79 |
| AD cases | 6052 (57.8) | 76.3 (8.2) | 47.54 | 39.29 | 4.23 | 6.08 | 2.46 | 0.4 |
| **Replication – (imputed SNP arrays)** |  |  |  |  |  |  |  |  |
| Controls | 10539 (59.4) | 76.7 (8.5) | 60.98 | 22.01 | 2.07 | 12.11 | 2.18 | 0.65 |
| AD cases | 11092 (60.5) | 73.3 (9.3) | 32.83 | 44.37 | 16.21 | 3.69 | 2.79 | 0.1 |

As a replication sample, we mega-analyzed 34 cohorts, each corresponding to a specific SNP array applied to an AD case/control dataset [3,17–25]. Some of these cohorts correspond to the same AD study but individuals were genotyped on different platforms. These cohorts are heterogenous in terms of age reported and are extensively described elsewhere [3,13] (**Tables 1, S2**). When multiple ages were available for a given subject, the order of priority for which age to use was AAO then age at examination then age at death in affected individuals, and age at death then age at last examination in control participants [13]. We removed any duplicated individuals across these cohorts and the discovery sample.

### Genetic quality control

For each cohort included in our analysis, we first determined the ancestry of each individual with SNPWeights v2.1[28] using reference populations from the 1000 Genomes Consortium [29]. Prior to ancestry determination, variants were filtered based on genotyping rate (< 95%), MAF < 1% and Hardy-Weinberg equilibrium (HWE) in controls (p < 10^−6^). By applying an ancestry percentage cut-off > 75%, the samples were stratified into five super populations: South-Asians, East-Asians, Americans, Africans and Europeans, and an Admixed group composed of individuals not passing cut-off in any single ancestry. Since most individuals were Europeans and to avoid spurious associations, we focused on European ancestry individuals.

Carriers of known pathogenic mutations on *APP, PSEN1, PSEN2* and *MAPT* were excluded from our analysis. Discordant pathology cases, defined as any clinically diagnosed AD individual with Braak stage below III or neuritic plaques level below moderate, were excluded from our analysis.

The joint called set of exome variants in the ADSP WES is composed of 1,524,414 SNPs [6,16]. We restricted downstream analysis to these variants, meaning that variants called only in ADSP WGS or AMP-AD were not included. To remove potential sequencing artefacts, we applied several quality control (QC) steps to each dataset. First, SNPs were checked for consistency with the Haplotype reference consortium (HRC) panel [30]. This check included flipping SNPs reported on incorrect strand and excluding SNPs with more than 10 % MAF difference with the HRC panel. Second, we removed SNPs that deviated from HWE in controls (p < 10^−6^) or that had a genotyping rate below 95%. Third, we removed any variants which had a flag different than PASS in gnomADv3. [31]. Following these QC steps 905,341 variants remained. For analysis, we considered 124,679 variants with minor allele count above 10, to ensure a minimum number of carriers.

In each cohort of the replication sample, SNPs with less than 95% genotyping rate or deviating from HWE in controls (p < 10^−6^) were excluded. Then, we used the gnomAD database [31] to filter out SNPs that met one of the following criteria: (i) located in low complexity region, (ii) located within common structural variants (MAF > 1%), (iii) multiallelic SNPs with MAF > 1 % for at least two alternate alleles, (iv) located within a common Ins/Del (insertion/deletion), (v) having any flag different than PASS in gnomAD, (vi) having potential probe polymorphisms [32]. The latter are defined as SNPs for which the probe may have variable affinity due to the presence of other SNP(s) within 20 bp and with MAF > 1 %. Individuals with more than 5 % genotype missingness were excluded. Imputation was performed on the Michigan imputation server using the TOPMed reference panel [33,34]. Per cohort, only variants with sufficient imputation quality (r^2^ > 0.3) were included in the replication analysis (**Table S3**).

Identity-by-descent was run to determine the relatedness between all individuals using PLINKv1.9 [35]. In the discovery sample, we kept only one version of duplicated individuals and removed first degree relatives keeping AD relatives over controls; and when both had a concordant diagnosis, we kept the younger case or older control. In the replication sample, we removed any individuals already present in the discovery, and for duplicate subjects we kept the copy from the SNP array with the highest genome coverage.

On the subset of remaining individuals, we computed genetic principal components to account for population stratification [36] in both the discovery and replication samples, separately.

### Statistics, association models, and AD-age score

We considered four main models: logistic regression on AD diagnosis adjusted for age, logistic regression on AD diagnosis, linear regression on a score integrating case-control status and age, and multivariate Cox regression on AAO. When AAO was not available the first known age with AD diagnosis was used. Our analyses removed individuals younger than 60 and censored maximum age at 100. We considered controls below 60 as uninformative and cases below 60 as early onset AD potentially due to a causal mutation.

For the third model we defined the AD-age score as follow:

- log(1-weight(age)) - 0.5 for controls;
- -log(weight(age)) + 0.5 for cases.

The score was designed to abide by the following rules: cases and controls should be clearly separated (maximum value for controls -0.5 and minimum value for cases +0.5, ensuring that the minimum difference between cases/controls is greater than 1); younger cases should have higher scores compared to older cases, and older controls should have lower scores than younger ones. This ensured that younger cases and older controls were at opposite extremes of the score spectrum and assumed these individuals influenced genetic associations the most.

We defined two *weight(age)* functions:

A. a linear definition: weight(age) = (age-59.5)/(100.5-59.5);
B. a piecewise continuous definition:
  ○ 60 and below: weight(age) = 5/320;
  ○ >60 to 65: weight(age) = (age-55)/320;
  ○ >65 to 75: weight(age) = 4*(age-55)/320 - 3/320;
  ○ >75 to 80: weight(age) = 10*(age-55)/320 - 15/320;
  ○ >80 to 90: weight(age) = 16*(age-55)/320 - 30/320;
  ○ >90 to 100: weight(age) = 6*(age-55)/320 + 5/320.

(A) corresponds to a linear effect of age between 60 and 100 and (B) accounts for the changes in AD prevalence slope in this age range [8].

For the analysis of exome-wide data, all models had two subversions: (1) adjusted for sex and 10 first principal components of population structure and (2) additionally adjusted for *APOE* ε2 and *APOE* ε4 alleles.

The associations for the first three models were estimated with PLINKv2.0 [37] using the *–glm* flag, which performs a logistic regression for case/control phenotype and a linear regression for quantitative phenotype. The Cox regression associations were estimated with *gwasurvir* [38].

We calculated the number of independent variants with PLINKv1.9 [35] (option –*indep-pairwise* 1000 50 0.1), which identified 87,034 linkage disequilibrium blocks covering the 124,679 considered variants. Thus, the exome-wide threshold was set at p < 5×10^−7^ (0.05/87034, Bonferroni correction) and the suggestive threshold at p < 1×10^−5^ (1/87304). A 1Mb region around the *APOE* locus was excluded from the reported results due to its well-established association with AD. We did not correct for the number of tested models due to their high correlation (cf. Results), nor for the two versions of adjustment (*APOE* ε2 and *APOE* ε4 alleles adjusted or not), as in Bis et al. [16], since these were similarly highly correlated.

Thirty-one variants passing the suggestive threshold in the discovery were evaluated in the replication sample. We disentangled spurious and true associations based on their associations in the replication dataset. SNVs with discordant direction of effect were considered to be spurious associations. Variants which had a concordant direction of effect and p < 1.6×10^−3^ (0.05/31, Bonferroni correction) for at least one model were considered significant, while those with p < 0.05 were considered to replicate nominally.

For more robust and powerful inference with the AD-age score, which is not normally distributed, we performed bootstrapping (100 resamplings) consistent with what was done in power simulations. To limit the computational burden, we only computed the bootstrap-based inference for the set of replicated variants, which allowed us to compare the significance of the linear regression on AD-age score with the Cox regression for true associations. Last, we performed a fixed-effect meta-analysis using the *metafor* package in R [39] to estimate the significance of the replicated variants in the combined discovery and replication samples.

### Gene and variant annotations

Each variant consequence was annotated with the Ensembl Variant Effect Predictor toolset [40]. Non-synonymous variants, such as missense or frameshift variants, may lead to loss or gain of function that may affect the enzymatic activity, stability, and/or interaction properties at the protein level. Synonymous variants, by contrast, do not typically directly affect protein function; however, they can influence protein expression both at the transcriptional and translational level [41].

To disentangle the role of the synonymous common variants as potential expression quantitative trait loci (eQTL), we queried the largest brain *cis*-eQTL meta-analysis which included 1,433 post-mortem brain samples from the AMP-AD and CommonMind Consortium [42].

Lastly, for mapped genes harboring significant variants, we queried the AMP-AD fixed-effect meta-analysis of gene differential expression between AD and control individuals across brain tissues [23,25,27].

## RESULTS

### Age-informed AD risk estimation increases power for genetic association testing

Power outcomes for specific illustrations of simulation analyses, considering a range of age-related risk effect estimates, are presented in **Figures 1, S1**. An overview of power differences between different association models for all simulations’ conditions, varying the AD risk associated with age, is provided in **Figure S2**. In simulations where the mean age of cases was younger than in controls, adjustment for age in logistic regression analyses compared to not adjusting for age led to critical power loss (**Figure 1**), amounting to as much as 90 % power loss in some conditions (**Figure S2 A-D**). The AD-age score model performed best overall across all four models, displaying power increases regardless of age differences between cases and controls, particularly for the estimated age effect on AD status [12] (**Figures 1, S2**). Power gain of the AD-age score with regard to logistic regression not adjusted for age was on average 10 %, up to 20 % in some scenarios (**Figure S2 C-D**). The Cox regression on AAO performed similarly as unadjusted logistic regression and in some scenarios performed better (**Figure 1 D-F**). When cases and controls were age-matched, the Cox regression displayed power losses for some conditions (**Figure S2 E-F**). Power gain of the AD-age score with regard to Cox regression was on average 8-10 %, reaching above 20 % in some scenarios (**Figure S2 G-H**). In any considered scenario, the AD-age score never decreased power in comparison to any other model (**Figure S2**).

**Figure 1.**
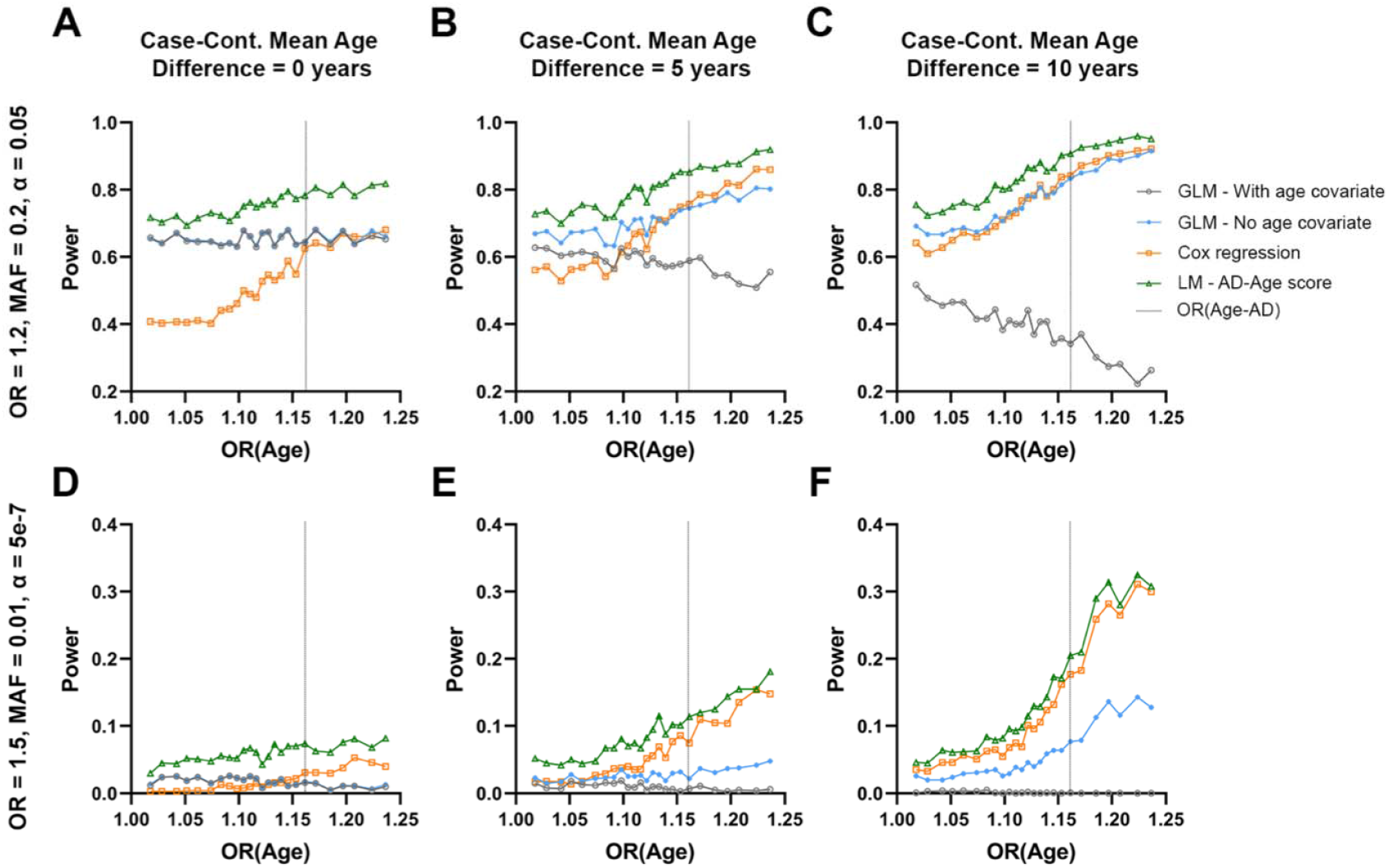
Power of different association models for two specific simulation outcomes. **A-C)** A common variant with moderate effect size, evaluated in 1000 cases and 1000 controls at a significance level of α = 0.05, mimicking the condition of common AD cohorts genotyped on SNP arrays. **D-F)** An uncommon variant with large effect size, evaluated in 5000 cases and 5000 controls at a significance level of α = 5×10^−7^, mimicking the condition of ADSP WES which allows exploration of uncommon and rare variant associations. Panels show power on the y-axis and age-related effect estimates on the x-axis. Outcomes for four models are shown (cf. legend) and the age-related effect estimate for AD [OR(Age-AD)] is marked by a vertical grey dotted line. From left to right, panels show simulation results for increasing mean age difference between cases and controls (cases being younger than controls where applicable).

**Figure 2.**
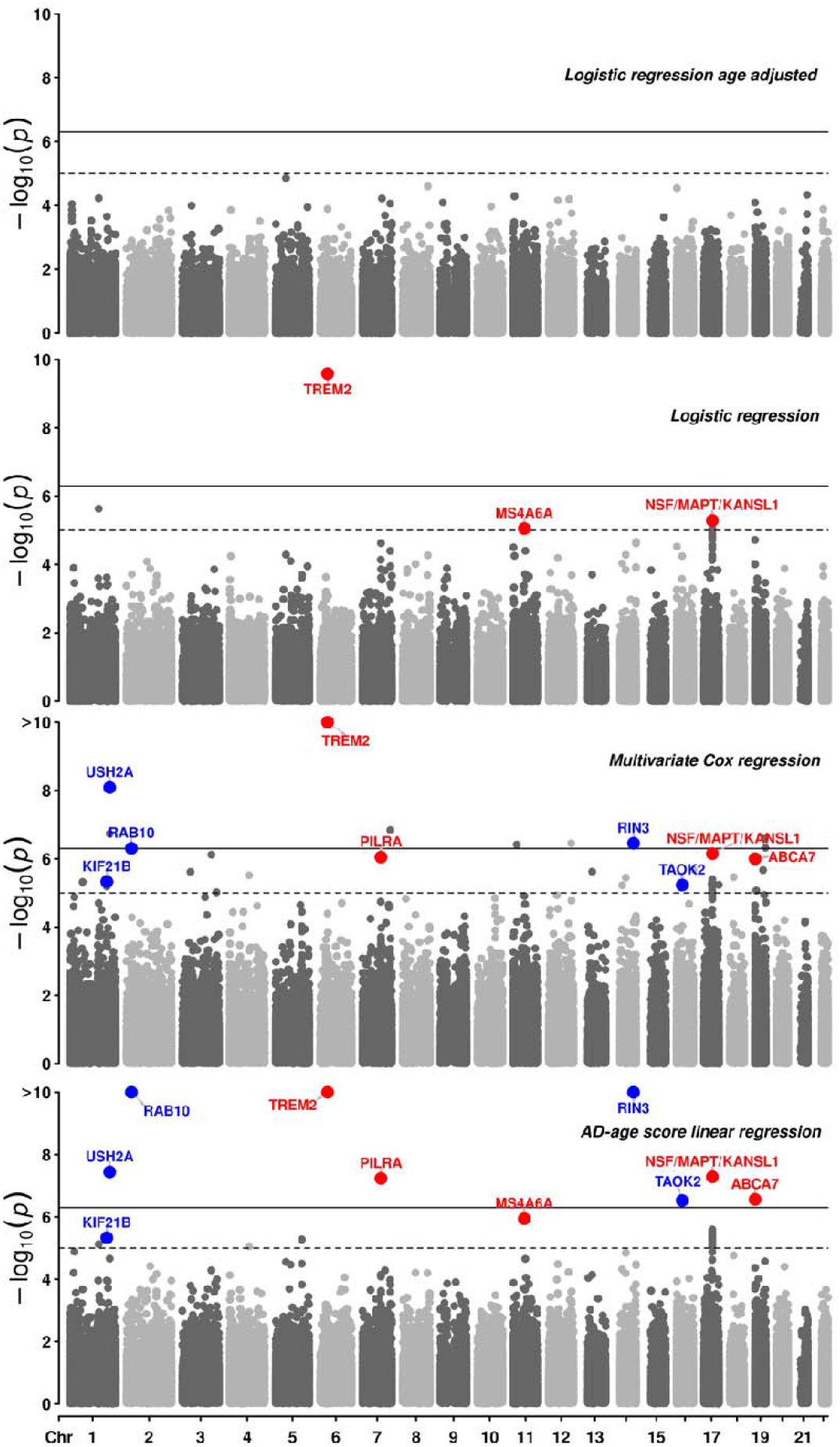
Manhattan plots of exome wide associations in the four main models excluding the *APOE* region. The age adjusted logistic regression has no suggestive association (dashed line, p < 1×10^−5^). The main causal variant on *TREM2* is exome wide significant (solid line, p < 5×10^−7^) in the other three models. Among suggestive associations, (i) known AD associations are in red, (ii) novel associations which replicate (p < 0.05) in an independent dataset are in blue (cf **Table 3**). Colored dots were bootstrapped in the AD-age score model (see Methods). The minimum p-value from the adjustment models for each main model is displayed as in [16].

### Exome-wide association

Exome-wide association with AD in the discovery sample for all four models and their subversions are shown in (**Figures S3-S6**). QQ plots for each exome-wide association show no inflation (λ < 1.1), except for the Cox regression adjusted for *APOE* ε2 and *APOE* ε4 allele dosages (λ = 1.19) (**Table S4, Figures S7-S10**). The logistic regression adjusted for age showed no associations above the suggestive threshold outside of the *APOE* region (**Figure S3**). Across the three other models a total of 31 variants passed suggestive significance, including 5 known AD risk loci [16]. The parameter estimate of these models: (i) OR (odd ratio) for logistic regression, (ii) exp(β) for the linear regression, and (iii) 1/HR (hazard ratio) for the Cox regression were found to be highly correlated (**Figure S11**), with (i-ii) Pearson correlation: r^2^= 0.80 (p = 3×10^−12^), (i-iii) r^2^ = 0.84 (p = 4×10^−14^), and (ii-iii) r^2^ = 0.97 (p < 2×10^−16^). The known *TREM2* missense single nucleotide variant (SNV) (rs75932628) was exome-wide significant in the three models. Other known associations included synonymous SNVs on *PILRA* (rs2405442), *MS4A6A* (rs12453), *NSF* (rs199533, lead SNV of a locus also encompassing *MAPT* and *KANSL1*), and a frameshift deletion on *ABCA7* (rs547447016) (**Figure 1, Tables 2, 3**). The association on *PILRA* was exome-wide significant in the AD-age score linear regression and suggestive in the Cox regression but did not reach the suggestive threshold in the logistic regression. Similarly, the association on *ABCA7* was suggestive in both AD-age score and Cox regressions, but not in the logistic regression. On the contrary, the association on *MS4A6A* was suggestive in the logistic regression and in the AD-age score and just below significance in the Cox regression. The association on *NSF/MAPT/KANSL1* was suggestive in all three models. In addition to these 5 known exonic associations, associations on 26 other exonic loci were at least suggestive in one of the three models **(Table S5**). Logistic regression (**Figure S4**) produced one spurious association on *ETV3L*, the AD-age score linear regression led to three spurious associations on *TACR3, PCDHA7*, and the one on *ETV3L*, while the Cox regression (**Figure S5**) had 16 spurious associations including the one on *TACR3*. The logistic regression model showed no novel suggestive association. The AD-age score linear regression, prior to bootstrap (**Figure S6**), produced two novel suggestive-level associations: one *USH2A* missense SNV (rs111033333) and one *RIN3* missense SNV (rs150221413), which replicated nominally. The Cox regression produced several exome-wide significant associations in the discovery with concordant direction of effect in the replication including *NAV2* (rs11828836), *RAB10* (rs149622307), and the *USH2A* and *RIN3* associations, also found in the AD-age score linear regression. Among suggestive associations in the Cox regression, two significantly replicated: *RAB10* synonymous SNV (rs149622307) and *TAOK2* synonymous SNV (rs4077410); and three nominally replicated: *KIF21B* synonymous SNV (rs2297911), and the previous missenses on *USH2A* and *RIN3. NAV2* synonymous SNV (rs11828836) did not reach nominal significance (p = 0.17), but it was imputed with sufficient quality in only 9,235 individuals (less than 50% of imputed individuals). *CDKL1* intronic SNV (rs61981931) did not reach nominal significance (p = 0.09).

**Table 2.** Main association results. Effect corresponds to OR (odds ratio) for logistic regression on AD status not adjusted by age (LogReg), exp(β) for linear regression on AD-age score (LinReg), and 1/HR (hazard ratio) for multivariate Cox regression on age-at-onset (CoxReg). Correlation between these measures is high for suggestive associations as shown on **Figure S11**. P: p-value. m: model subversion. Subversion codes are: (1) adjusted for sex and 10 first principal components of population structure and (2) additionally adjusted for *APOE* ε2 and *APOE* ε4 alleles. Two types of weighted AD-age score were used with (A) corresponding to a linear effect of age between 60 and 100 and (B) accounting for the changes in AD prevalence slope in this age range [8].

| SNP (hg19) / Gene | Discovery |  |  |  |  |  |  |  |  | Replication |  |  |  |  |  |  |  |  |
| --- | --- | --- | --- | --- | --- | --- | --- | --- | --- | --- | --- | --- | --- | --- | --- | --- | --- | --- |
|  | LogReg |  |  | LinReg |  |  | CoxReg |  |  | LogReg |  |  | LinReg |  |  | CoxReg |  |  |
| | OR | P | m | $\exp(\beta)$ | P | m | 1/HR | P | m | OR | P | m | $\exp(\beta)$ | P | m | 1/HR | P | m |
| 1:200959302:G:A / *KIF21B* | 0.87 | $2.10^{-4}$ | 2 | 0.90 | $5.10^{-6}$ | B2 | 0.89 | $5.10^{-6}$ | 2 | 0.96 | 0.13 | 1 | 0.96 | 0.01 | B1 | 0.96 | 0.02 | 2 |
| 1:216270469:G:A / *USH2A* | 9.12 | $4.10^{-3}$ | 2 | 6.76 | $4.10^{-8}$ | B1 | 4.07 | $8.10^{-9}$ | 2 | 1.58 | 0.14 | 1 | 1.70 | 0.04 | A1 | 1.33 | 0.12 | 1 |
| 2:26332640:T:C / *RAB10* | 17.4 | 0.06 | 1 | 10.46 | $2.10^{-15}$ | B1 | 4.92 | $5.10^{-7}$ | 1 | 4.50 | 0.05 | 1 | 5.03 | $2.10^{-3}$ | B1 | 2.69 | $6.10^{-4}$ | 1 |
| 6:41129252:C:T / *TREM2* | 4.83 | $3.10^{-10}$ | 1 | 3.22 | $2.10^{-27}$ | A1 | 2.58 | $1.10^{-23}$ | 1 | 2.32 | $2.10^{-9}$ | 1 | 2.69 | $1.10^{-14}$ | A1 | 1.95 | $2.10^{-18}$ | 2 |
| 7:99971313:T:C / *PILRA* | 0.88 | $2.10^{-5}$ | 1 | 0.87 | $6.10^{-8}$ | A2 | 0.90 | $9.10^{-7}$ | 2 | 0.92 | $6.10^{-5}$ | 1 | 0.90 | $2.10^{-7}$ | B1 | 0.93 | $5.10^{-7}$ | 1 |
| 11:59945745:T:C / *MS4A6A* | 0.88 | $9.10^{-6}$ | 1 | 0.91 | $1.10^{-6}$ | B1 | 0.92 | $1.10^{-5}$ | 1 | 0.89 | $1.10^{-8}$ | 1 | 0.89 | $3.10^{-12}$ | A1 | 0.93 | $2.10^{-8}$ | 1 |
| 14:93022240:G:T / *RIN3* | 16.3 | $7.10^{-3}$ | 2 | 6.54 | $6.10^{-11}$ | A2 | 3.46 | $4.10^{-7}$ | 2 | 1.95 | 0.04 | 2 | 1.69 | 0.02 | A2 | 1.59 | 0.01 | 2 |
| 16:29998200:A:G / *TAOK2* | 1.12 | $6.10^{-5}$ | 1 | 1.08 | $3.10^{-7}$ | A1 | 1.09 | $6.10^{-6}$ | 2 | 1.04 | 0.07 | 2 | 1.05 | $1.10^{-3}$ | B1 | 1.05 | $4.10^{-4}$ | 2 |
| 17:44828931:G:A / *NSF/MAPT/KANSL1* | 0.85 | $5.10^{-6}$ | 2 | 0.89 | $5.10^{-8}$ | B2 | 0.89 | $7.10^{-7}$ | 2 | 0.97 | 0.20 | 2 | 0.97 | 0.06 | B2 | 0.98 | 0.18 | 2 |
| 19:1047507:AGGAGCAG:A / *ABCA7* | 3.36 | $1.10^{-4}$ | 2 | 2.18 | $3.10^{-7}$ | A1 | 1.94 | $1.10^{-6}$ | 1 | 1.36 | 0.12 | 1 | 1.33 | 0.07 | B2 | 1.22 | 0.13 | 2 |

**Table 3.** Sample sizes, minor allele frequency and imputation quality for the identified variants. MAF: Minor allele frequency; R-square (Rsq): Imputation quality.

| Gene(s) | RS id | Consequence | SNP (hg19) | Discovery |  | Replication |  |  |
| --- | --- | --- | --- | --- | --- | --- | --- | --- |
|  |  |  |  | N | MAF | N | MAF | Rsq |
| *KIF21B* | rs2297911 | synonymous | 1:200959302:G:A | 11006 | 0.17391 | 21631 | 0.1769 | 1 |
| *USH2A* | rs111033333 | missense | 1:216270469:G:A | 11126 | 0.00085 | 19544 | 0.00132 | 0.81 |
| *RAB10* | rs149622307 | synonymous | 2:26332640:T:C | 11057 | 0.00045 | 9833 | 0.00076 | 0.85 |
| *TREM2* | rs75932628 | missense | 6:41129252:C:T | 11076 | 0.00591 | 21176 | 0.00606 | 0.93 |
| *PILRA* | rs2405442 | synonymous | 7:99971313:T:C | 11022 | 0.29836 | 21631 | 0.30567 | 0.94 |
| *MS4A6A* | rs12453 | synonymous | 11:59945745:T:C | 11114 | 0.3941 | 21481 | 0.39015 | 0.99 |
| *RIN3* | rs150221413 | missense | 14:93022240:G:T | 11020 | 0.00082 | 17652 | 0.00131 | 0.8 |
| *TAOK2* | rs4077410 | synonymous | 16:29998200:A:G | 11063 | 0.47966 | 21631 | 0.48195 | 0.94 |
| *NSF/MAPT/ KANSL1* | rs199533 | synonymous | 17:44828931:G:A | 11094 | 0.20367 | 21631 | 0.19931 | 0.99 |
| *ABCA7* | rs547447016 | frameshift | 19:1047507:AGGAGCAG:A | 11006 | 0.00313 | 18356 | 0.00311 | 0.88 |

For the set of replicated variants (**Table 2**), we meta-analyzed the discovery and independent replication results. Seven out of the ten exonic variants were most significant in the linear regression on the AD-age score, while only two performed best in the Cox regression, those on *KIF21B* and *TAOK2*, and one in the logistic regression, on *MS4A6A* (**Figure S12**). After meta-analysis, the variants located on *RAB10, TREM2, PILRA, MS4A6A*, and *RIN3* were exome-wide significant (p <5×10^−7^) (**Table S6**).

### Functional annotation

Among the mapped genes (**Table 3**), the synonymous variants on *PILRA* and *KANSL1* were significantly associated with the expression of their respective mapped gene (false discovery rate (FDR) corrected). At the nominal significance level, *TAOK2* and *KIF21B* synonymous variants were also associated with the expression of their respective genes. Among nearby genes with FDR-significant eQTL association, *PVRIG* was the strongest association at the *PILRA* locus, *KANSL1-AS1* at the *NSF/MAPT/KANSL1* locus, and *INO80E* at the *TAOK2* locus (**Table S6**). In the meta-analysis of differential gene expression across brain tissues in AMP-AD: *TREM2, KANSL1, RAB10, MS4A6A*, and *RIN3* were found to be significantly upregulated in AD compared to control individuals, while *TAOK2* was significantly downregulated (reported associations were FDR-significant, **Table S7**).

## DISCUSSION

In the AD data simulation, we showed that incorrectly adjusting for age led to critical power loss and that weighting the known effect of age on AD risk in the phenotype increased statistical power for variant discovery. Testing these models on real AD data confirmed our simulation observations and enabled the discovery of novel variants modulating AD risk.

### Previous literature

The main prior AD WES study aimed to address the age adjustment conundrum in the ADSP WES data by implementing three different logistic regression models: the main one being unadjusted for age, while the other two were age adjusted [7]. However, given that cases were on average younger than controls, the age adjustment was in the opposite direction of the true age effect on AD risk. It is perhaps unsurprising, therefore, that there were no replicated findings from the two age-adjusted models (only associations from the main age-unadjusted model in the ADSP discovery were replicated) [7].

An alternative approach has been to use Cox regression on AAO for improved power compared to logistic regression that only considers case-control status. Cox regression has proven successful in predicting an individual’s AD conversion risk by calculating a polygenic hazard score [43,44]. However, it needs to abide by several assumptions, including proportional hazards across age. Several studies have shown that Cox regression performs better than logistic regression on case-control data when AAO is available [45,46], but it has not been applied to the ADSP WES data. Cox regression was previously applied to AD GWA, using genotype-imputed data overlapping partially with the ADSP sample used here, and led to the discovery of novel associations [47]. Alternative approaches have been proposed when Cox regression’s assumptions are violated as in AD GWA, including age stratification [48] and generalized Cox regression [49]. Our proposed AD-age score offers additional flexibility without these assumptions and it can accommodate age information other than AAO such as age-at-study and age-at-death. Unlike Cox regression models, the AD-age score can be flexibly incorporated as a quantitative outcome into conventional tools (e.g. PLINK) for GWAS and new methods (e.g. BOLT-LMM, SAIGE) for analysis of large/biobank scale genetic data with related samples. Additionally, the linear and logistic regressions are faster than Cox regression and thus more advantageous for larger datasets [45].

Oversampling cases with early AAO and controls with late censoring time for exome sequencing is an efficient design because it directs limited study resources towards subjects that are most useful for discovering the genetic associations of AD in the original cohorts [50,51]. We proposed the AD-age score for improved power in the discovery stage, and validated the findings using an independent replication sample. Although the hypothesis testing is appropriate in the discovery stage with extreme sampling, it is worth noting that the estimated genetic effect / odds ratio may not represent that in the whole population [52]. To obtain unbiased genetic effect estimations of AD risk in the whole population, it may be advisable to turn to more advanced methods that can explicitly address the biased sampling design (e.g., [50,53]).

### Potential disease mechanisms

The novel variants identified through our exome-wide association, with the exception of the *USH2A* SNV, are located on genes previously linked to AD, re-enforcing our confidence in these associations.

Our main finding is a rare variant on *RAB10* passing the exome-wide threshold in discovery and surviving Bonferroni correction in the replication. RAB proteins are key regulators of vesicular trafficking and play a major role in the endolysosomal and retromer pathways known to be linked to AD [54]. Another rare *RAB10* SNV was shown to segregate with AD resilience in pedigrees at risk for AD and *RAB10* was shown to be upregulated in AD brains [55], a finding corroborated in our study. *RAB10* knockdown significantly decreased Aβ_42_ and Aβ_42_/Aβ_40_ ratio in neuroblastoma cells [55]. Silencing of *RAB10* decreased β-amyloid peptides (Aβ) and increased soluble ectodomain of APP β (sAPPβ) [56], supporting a role of *RAB10* in either γ-secretase cleavage of APP and the degradation of Aβ. Moreover, phosphorylated Rab10 was prominent in neurofibrillary tangles in the hippocampus of AD individuals but scarce in controls [57]. Mechanistically, the JNK-interactin protein 1 (JIP1), mediates the anterograde transport of Rab10-positive cargo to axonal tips which promotes axonal growth and is critical for proper neuronal function [58]. JIP1 also regulates anterograde and retrograde transport of APP along axons [59]. These molecular mechanisms suggest that Rab10 could play a role in APP trafficking along axons.

Additionally, our exome-wide analysis identified a missense variant on Rab interactor 3 (*RIN3*). Common variants in a locus near *RIN3* and *SLC24A4*, were reported to be associated with AD susceptibility [2]. Increased *RIN3* expression in *APP/PS1* mouse models was shown to correlate with endosomal dysfunction and altered axonal trafficking and processing of *APP* [60]. For these reasons, the Rab related proteins involved in the endolysosomal and retromer pathways have been considered as promising therapeutic targets for AD [54].

Two common exonic variants, on *TAOK2* and *KIF21B*, were identified as suggestive in our discovery analysis and replicated (Bonferroni corrected and nominally, respectively). Previous AD GWAS summary statistics show a concordant direction of effect with our analysis [2,3] with the SNVs p-values on *TAOK2* and *KIF21B* in those studies equal to 0.05 and 10^−5^, respectively. *TAOK2* was shown to be phosphorylated in AD and frontotemporal lobar degeneration brains. Its expression was colocalized with tangles and its inhibition reduced tau phosphorylation [61]. Further, *KIF21B* is involved in neuronal and synaptic signaling and increased *KIF21B* expression levels were associated with more severe AD pathology [62].

### Limitation

For common synonymous variants, the regulated gene and true causal variant remain uncertain because our study focused on exomes and we cannot perform a genome-wide colocalization analysis. The causal variant may be intergenic and in linkage disequilibrium with a common synonymous variant identified in our analysis. Thus, future genome-wide studies are warranted to help disentangle which nearby genes are regulated, notably for the novel common loci encompassing *KIF21B* and *TAOK2*.

### Conclusion

Correctly accounting for the risk-increasing effect of age on AD is an efficient means of increasing statistical power. Thus, our AD-age score should prove useful in future AD genetic association studies to enable the discovery of additional novel variants.

## Data Availability

All samples were available from publicly released AD-related cohorts, with phenotype and genotype ascertainment described elsewhere [See references 3,6,17-27,13].
Notably from NIAGADS and dbgap repositories.

## ACKNOWLEDGMENTS

Funding for this study was provided by the Iqbal Farrukh & Asad Jamal Fund, the National Institutes of Health (grants AG066206, AG060747 and AG047366), and the Alzheimer’s Association (AARF-20-683984, M.E.B).

The Alzheimer’s Disease Sequencing Project (ADSP) is comprised of two Alzheimer’s Disease (AD) genetics consortia and three National Human Genome Research Institute (NHGRI) funded Large Scale Sequencing and Analysis Centers (LSAC). The two AD genetics consortia are the Alzheimer’s Disease Genetics Consortium (ADGC) funded by NIA (U01 AG032984), and the Cohorts for Heart and Aging Research in Genomic Epidemiology (CHARGE) funded by NIA (R01 AG033193), the National Heart, Lung, and Blood Institute (NHLBI), other National Institute of Health (NIH) institutes and other foreign governmental and non-governmental organizations. The Discovery Phase analysis of sequence data is supported through UF1AG047133 (to Drs. Schellenberg, Farrer, Pericak-Vance, Mayeux, and Haines); U01AG049505 to Dr. Seshadri; U01AG049506 to Dr. Boerwinkle; U01AG049507 to Dr. Wijsman; and U01AG049508 to Dr. Goate and the Discovery Extension Phase analysis is supported through U01AG052411 to Dr. Goate, U01AG052410 to Dr. Pericak-Vance and U01 AG052409 to Drs. Seshadri and Fornage.

The ADGC cohorts include: Adult Changes in Thought (ACT) (UO1 AG006781, UO1 HG004610, UO1 HG006375, U01 HG008657), the Alzheimer’s Disease Centers (ADC) (P30 AG019610, P30 AG013846, P50 AG008702, P50 AG025688, P50 AG047266, P30 AG010133, P50 AG005146, P50 AG005134, P50 AG016574, P50 AG005138, P30 AG008051, P30 AG013854, P30 AG008017, P30 AG010161, P50 AG047366, P30 AG010129, P50 AG016573, P50 AG016570, P50 AG005131, P50 AG023501, P30 AG035982, P30 AG028383, P30 AG010124, P50 AG005133, P50 AG005142, P30 AG012300, P50 AG005136, P50 AG033514, P50 AG005681, and P50 AG047270), the Chicago Health and Aging Project (CHAP) (R01 AG11101, RC4 AG039085, K23 AG030944), Indianapolis Ibadan (R01 AG009956, P30 AG010133), the Memory and Aging Project (MAP) (R01 AG17917), Mayo Clinic (MAYO) (R01 AG032990, U01 AG046139, R01 NS080820, RF1 AG051504, P50 AG016574), Mayo Parkinson’s Disease controls (NS039764, NS071674, 5RC2HG005605), University of Miami (R01 AG027944, R01 AG028786, R01 AG019085, IIRG09133827, A2011048), the Multi-Institutional Research in Alzheimer’s Genetic Epidemiology Study (MIRAGE) (R01 AG09029, R01 AG025259), the National Cell Repository for Alzheimer’s Disease (NCRAD) (U24 AG21886), the National Institute on Aging Late Onset Alzheimer’s Disease Family Study (NIA-LOAD) (R01 AG041797), the Religious Orders Study (ROS) (P30 AG10161, R01 AG15819), the Texas Alzheimer’s Research and Care Consortium (TARCC) (funded by the Darrell K Royal Texas Alzheimer’s Initiative), Vanderbilt University/Case Western Reserve University (VAN/CWRU) (R01 AG019757, R01 AG021547, R01 AG027944, R01 AG028786, P01 NS026630, and Alzheimer’s Association), the Washington Heights-Inwood Columbia Aging Project (WHICAP) (RF1 AG054023), the University of Washington Families (VA Research Merit Grant, NIA: P50AG005136, R01AG041797, NINDS: R01NS069719), the Columbia University Hispanic Estudio Familiar de Influencia Genetica de Alzheimer (EFIGA) (RF1 AG015473), the University of Toronto (UT) (funded by Wellcome Trust, Medical Research Council, Canadian Institutes of Health Research), and Genetic Differences (GD) (R01 AG007584). The CHARGE cohorts are supported in part by National Heart, Lung, and Blood Institute (NHLBI) infrastructure grant HL105756 (Psaty), RC2HL102419 (Boerwinkle) and the neurology working group is supported by the National Institute on Aging (NIA) R01 grant AG033193.

The CHARGE cohorts participating in the ADSP include the following: Austrian Stroke Prevention Study (ASPS), ASPS-Family study, and the Prospective Dementia Registry-Austria (ASPS/PRODEM-Aus), the Atherosclerosis Risk in Communities (ARIC) Study, the Cardiovascular Health Study (CHS), the Erasmus Rucphen Family Study (ERF), the Framingham Heart Study (FHS), and the Rotterdam Study (RS). ASPS is funded by the Austrian Science Fond (FWF) grant number P20545-P05 and P13180 and the Medical University of Graz. The ASPS-Fam is funded by the Austrian Science Fund (FWF) project I904), the EU Joint Programme - Neurodegenerative Disease Research (JPND) in frame of the BRIDGET project (Austria, Ministry of Science) and the Medical University of Graz and the Steiermärkische Krankenanstalten Gesellschaft. PRODEM-Austria is supported by the Austrian Research Promotion agency (FFG) (Project No. 827462) and by the Austrian National Bank (Anniversary Fund, project 15435. ARIC research is carried out as a collaborative study supported by NHLBI contracts (HHSN268201100005C, HHSN268201100006C, HHSN268201100007C, HHSN268201100008C, HHSN268201100009C, HHSN268201100010C, HHSN268201100011C, and HHSN268201100012C). Neurocognitive data in ARIC is collected by U01 2U01HL096812, 2U01HL096814, 2U01HL096899, 2U01HL096902, 2U01HL096917 from the NIH (NHLBI, NINDS, NIA and NIDCD), and with previous brain MRI examinations funded by R01-HL70825 from the NHLBI. CHS research was supported by contracts HHSN268201200036C, HHSN268200800007C, N01HC55222, N01HC85079, N01HC85080, N01HC85081, N01HC85082, N01HC85083, N01HC85086, and grants U01HL080295 and U01HL130114 from the NHLBI with additional contribution from the National Institute of Neurological Disorders and Stroke (NINDS). Additional support was provided by R01AG023629, R01AG15928, and R01AG20098 from the NIA. FHS research is supported by NHLBI contracts N01-HC-25195 and HHSN268201500001I. This study was also supported by additional grants from the NIA (R01s AG054076, AG049607 and AG033040 and NINDS (R01 NS017950). The ERF study as a part of EUROSPAN (European Special Populations Research Network) was supported by European Commission FP6 STRP grant number 018947 (LSHG-CT-2006-01947) and also received funding from the European Community’s Seventh Framework Programme (FP7/2007-2013)/grant agreement HEALTH-F4-2007-201413 by the European Commission under the programme “Quality of Life and Management of the Living Resources” of 5th Framework Programme (no. QLG2-CT-2002-01254). High-throughput analysis of the ERF data was supported by a joint grant from the Netherlands Organization for Scientific Research and the Russian Foundation for Basic Research (NWO-RFBR 047.017.043). The Rotterdam Study is funded by Erasmus Medical Center and Erasmus University, Rotterdam, the Netherlands Organization for Health Research and Development (ZonMw), the Research Institute for Diseases in the Elderly (RIDE), the Ministry of Education, Culture and Science, the Ministry for Health, Welfare and Sports, the European Commission (DG XII), and the municipality of Rotterdam. Genetic data sets are also supported by the Netherlands Organization of Scientific Research NWO Investments (175.010.2005.011, 911-03-012), the Genetic Laboratory of the Department of Internal Medicine, Erasmus MC, the Research Institute for Diseases in the Elderly (014-93-015; RIDE2), and the Netherlands Genomics Initiative (NGI)/Netherlands Organization for Scientific Research (NWO) Netherlands Consortium for Healthy Aging (NCHA), project 050-060-810. All studies are grateful to their participants, faculty and staff. The content of these manuscripts is solely the responsibility of the authors and does not necessarily represent the official views of the National Institutes of Health or the U.S. Department of Health and Human Services.

The four LSACs are: the Human Genome Sequencing Center at the Baylor College of Medicine (U54 HG003273), the Broad Institute Genome Center (U54HG003067), The American Genome Center at the Uniformed Services University of the Health Sciences (U01AG057659), and the Washington University Genome Institute (U54HG003079).

Biological samples and associated phenotypic data used in primary data analyses were stored at Study Investigators institutions, and at the National Cell Repository for Alzheimer’s Disease (NCRAD, U24AG021886) at Indiana University funded by NIA. Associated Phenotypic Data used in primary and secondary data analyses were provided by Study Investigators, the NIA funded Alzheimer’s Disease Centers (ADCs), and the National Alzheimer’s Coordinating Center (NACC, U01AG016976) and the National Institute on Aging Genetics of Alzheimer’s Disease Data Storage Site (NIAGADS, U24AG041689) at the University of Pennsylvania, funded by NIA This research was supported in part by the Intramural Research Program of the National Institutes of health, National Library of Medicine. Contributors to the Genetic Analysis Data included Study Investigators on projects that were individually funded by NIA, and other NIH institutes, and by private U.S. organizations, or foreign governmental or nongovernmental organizations.

The NACC database is funded by NIA/NIH Grant U01 AG016976. NACC data are contributed by the NIA-funded ADCs: P30 AG019610 (PI Eric Reiman, MD), P30 AG013846 (PI Neil Kowall, MD), P30 AG062428-01 (PI James Leverenz, MD) P50 AG008702 (PI Scott Small, MD), P50 AG025688 (PI Allan Levey, MD, PhD), P50 AG047266 (PI Todd Golde, MD, PhD), P30 AG010133 (PI Andrew Saykin, PsyD), P50 AG005146 (PI Marilyn Albert, PhD), P30 AG062421-01 (PI Bradley Hyman, MD, PhD), P30 AG062422-01 (PI Ronald Petersen, MD, PhD), P50 AG005138 (PI Mary Sano, PhD), P30 AG008051 (PI Thomas Wisniewski, MD), P30 AG013854 (PI Robert Vassar, PhD), P30 AG008017 (PI Jeffrey Kaye, MD), P30 AG010161 (PI David Bennett, MD), P50 AG047366 (PI Victor Henderson, MD, MS), P30 AG010129 (PI Charles DeCarli, MD), P50 AG016573 (PI Frank LaFerla, PhD), P30 AG062429-01(PI James Brewer, MD, PhD), P50 AG023501 (PI Bruce Miller, MD), P30 AG035982 (PI Russell Swerdlow, MD), P30 AG028383 (PI Linda Van Eldik, PhD), P30 AG053760 (PI Henry Paulson, MD, PhD), P30 AG010124 (PI John Trojanowski, MD, PhD), P50 AG005133 (PI Oscar Lopez, MD), P50 AG005142 (PI Helena Chui, MD), P30 AG012300 (PI Roger Rosenberg, MD), P30 AG049638 (PI Suzanne Craft, PhD), P50 AG005136 (PI Thomas Grabowski, MD), P30 AG062715-01 (PI Sanjay Asthana, MD, FRCP), P50 AG005681 (PI John Morris, MD), P50 AG047270 (PI Stephen Strittmatter, MD, PhD).

## Notes

### Competing Interest Statement

The authors have declared no competing interest.

### Author Declarations

The current study protocol was granted an exemption by the Stanford University institutional review board because the analyses were carried out on deidentified, off-the-shelf data; therefore, further informed consent was not required.

