## Supplementary Materials for "A novel age-informed approach for genetic association analysis in Alzheimer’s disease"

*Data used in preparation of this article were obtained from the Alzheimer's Disease Neuroimaging Initiative (ADNI) database (adni.loni.usc.edu). As such, the investigators within the ADNI contributed to the design and implementation of ADNI and/or provided data but did not participate in analysis or writing of this report. A complete listing of ADNI investigators can be found at:

http://adni.loni.usc.edu/wp-content/uploads/how_to_apply/ADNI_Acknowledgement_List.pdf

**Corresponding Author**

Yann Le Guen

Department of Neurology and Neurological Sciences – FIND lab

Stanford University

290 Jane Stanford Way, Stanford, CA, USA

**Figure S1. Power of different association models for two additional specific simulation outcomes.**

**Figure S2. Power differences between association models on simulated case-control data, considering the age-related risk effect estimate of Alzheimer's disease (OR 1.16).**

**Figure S3. Manhattan plots for the two model adjustments of the logistic regression adjusted by age.**

**Figure S4. Manhattan plots for the two model adjustments of the logistic regression not adjusted by age.**

**Figure S5. Manhattan plots for the two model adjustments of the multivariate Cox regression.**

**Figure S6. Manhattan plots for the two model adjustments on the two AD-age scores linear regression.**

**Figure S7. QQ plots of the logistic regression adjusted by age corresponding to Figure S3.**

**Figure S8. QQ plots of the standard logistic regression corresponding to Figure S4.**

**Figure S9. QQ plots of the multivariate Cox regression corresponding to Figure S5.**

**Figure S10. QQ plots of the linear regression on the AD-age score corresponding to Figure S6.**

**Figure S11. Comparison between exp(β), OR (odds ratio), 1/HR (hazard ratio) for associations suggestive in any models.**

**Figure S12. Comparison between -log(p) between logistic regression, linear regression on the AD-age score and multivariate Cox regression.**

**Table S1. Demographics per cohort in the discovery sample.**

**Table S2. Demographics per cohort in the replication sample.**

**Table S3. Number of individuals, minor allele frequency and imputation quality for the suggestive variants in the discovery outside of the *APOE* region.**

**Table S4. Lambda medians for each main model and model adjustments.**

**Table S5. All suggestive association results in the discovery outside of the *APOE* region.**

**Table S6. Meta-analysis of the replicated exonic associations.**

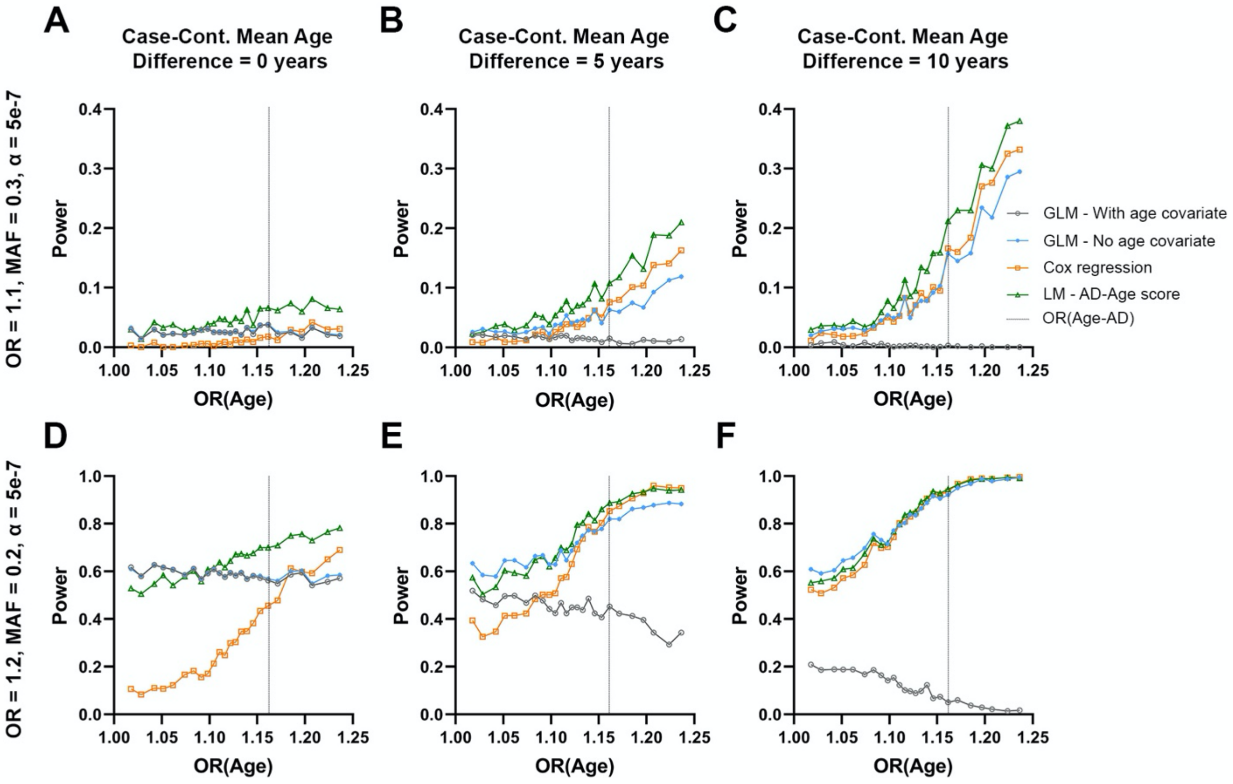


**Figure S1. Power of different association models for two additional specific simulation outcomes**. **A-C)** A highly common variant with small effect size, evaluated in 5000 cases and 5000 controls at a significance level of $\alpha = 5e^{-7}$. **D-F)** A common variant with moderate effect size, evaluated in 5000 cases and 5000 controls at a significance level of $\alpha = 5e^{-7}$. Panels show power on the y-axis and age-related effect estimates on the x-axis. Outcomes for four models are shown (cf. legend) and the age-related effect estimate for AD [OR(Age-AD)] is marked by a vertical grey dotted line. From left to right, panels show simulation results for increasing mean age differences between cases and controls (cases being younger where applicable). Note for **F)**, which mimics the ADSP WES study design, that for a common variant with moderate effect, there is no clear difference between logistic regression not adjusted for age, Cox regression, or the AD-age score. This is consistent with the expectation of ADSP's study design to increase statistical power by selecting for young cases versus old controls.

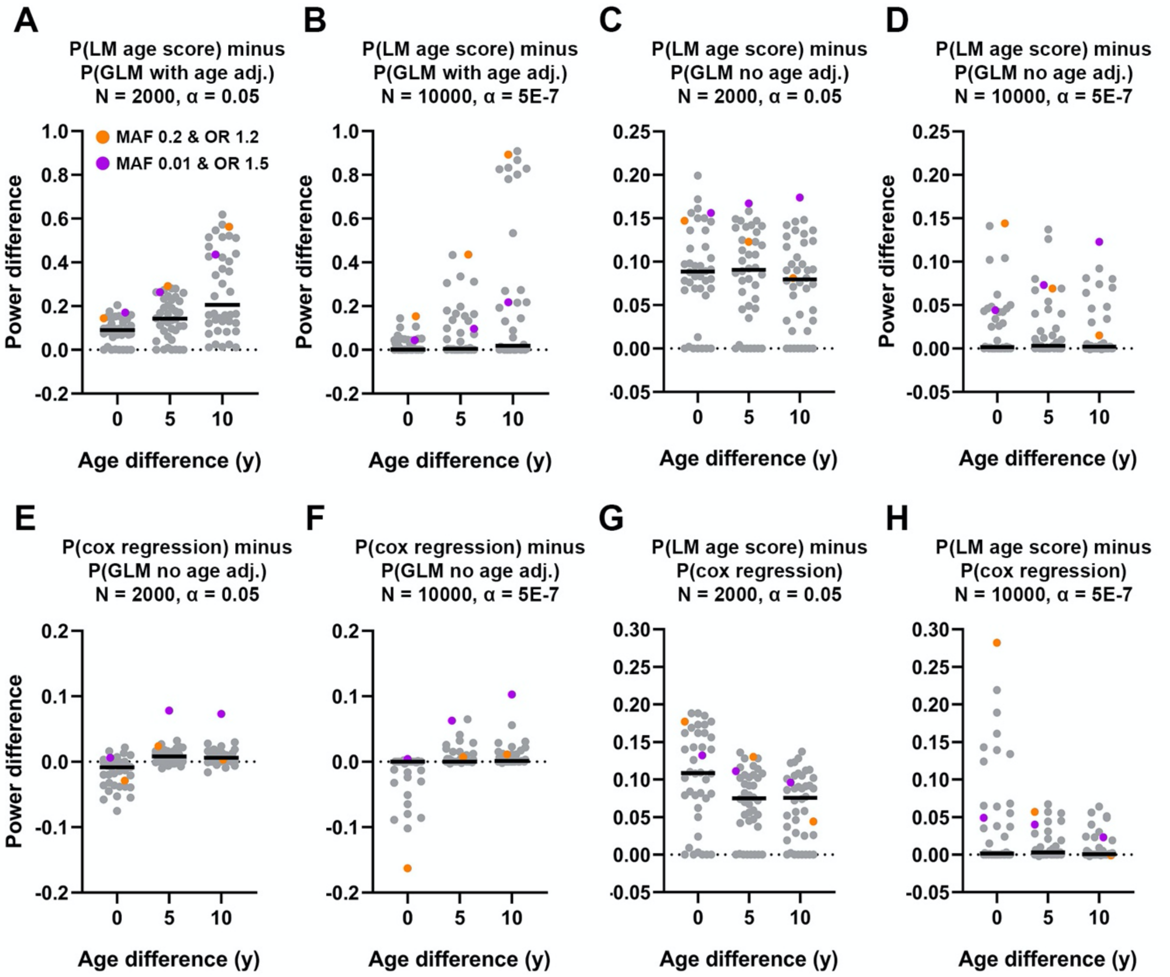


**Figure S2. Power differences between association models on simulated case-control data, considering the age-related risk effect estimate of Alzheimer's disease (OR 1.16).** Paired model comparisons and parameters are indicated on panel titles, where N = 2000 indicates 1000 cases and 1000 controls, and N = 10000 indicates 5000 cases and 5000 controls. Each panel displays power differences (y-axis) for each individual combination of simulated variant MAF and OR (single dot) and their averages (black line), stratified according to the mean age differences between cases and controls (x-axis). Orange and purple dots on each panel indicate power differences for conditions matching those presented in Figure 1. **A-B)** Critical power gain was observed when not adjusting for age in logistic regression analyses. **C-D)** There was on average 10% increase in power when using the AD-age score compared to logistic regression analyses not adjusting for age. This overall effect diminishes in a larger sample (N = 10000), but some conditions (e.g. purple dot) still display power gain. There was never any power loss. **E-F)** There was limited power gain when using the Cox regression and power loss was observed when there was no mean age difference between cases and controls. **G-H)** AD-age score always outperformed Cox regression.

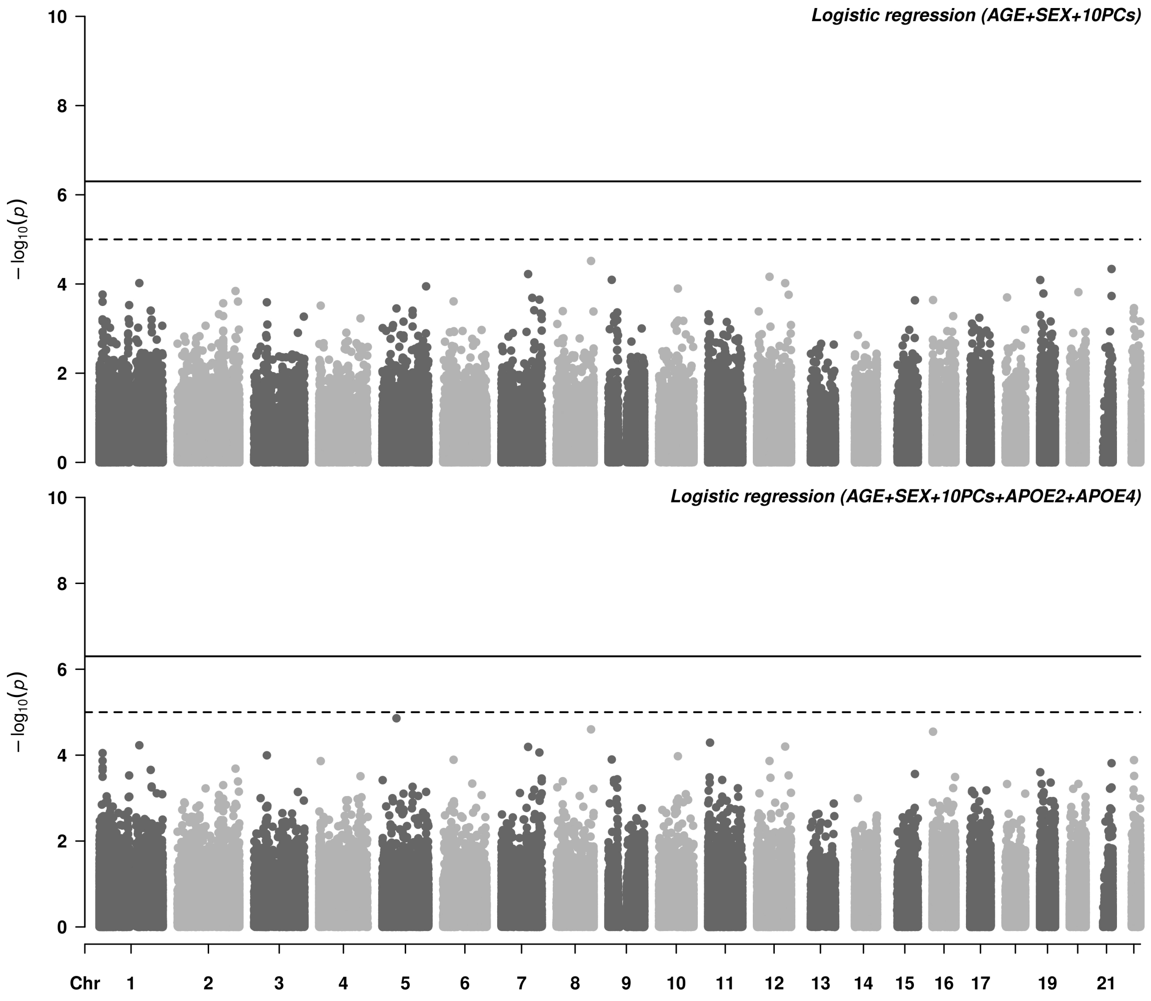


**Figure S3. Manhattan plots for the two model adjustments of the logistic regression adjusted by age.** The age adjusted logistic regression has no suggestive association (dashed line, $p < 1\times10^{-5}$).

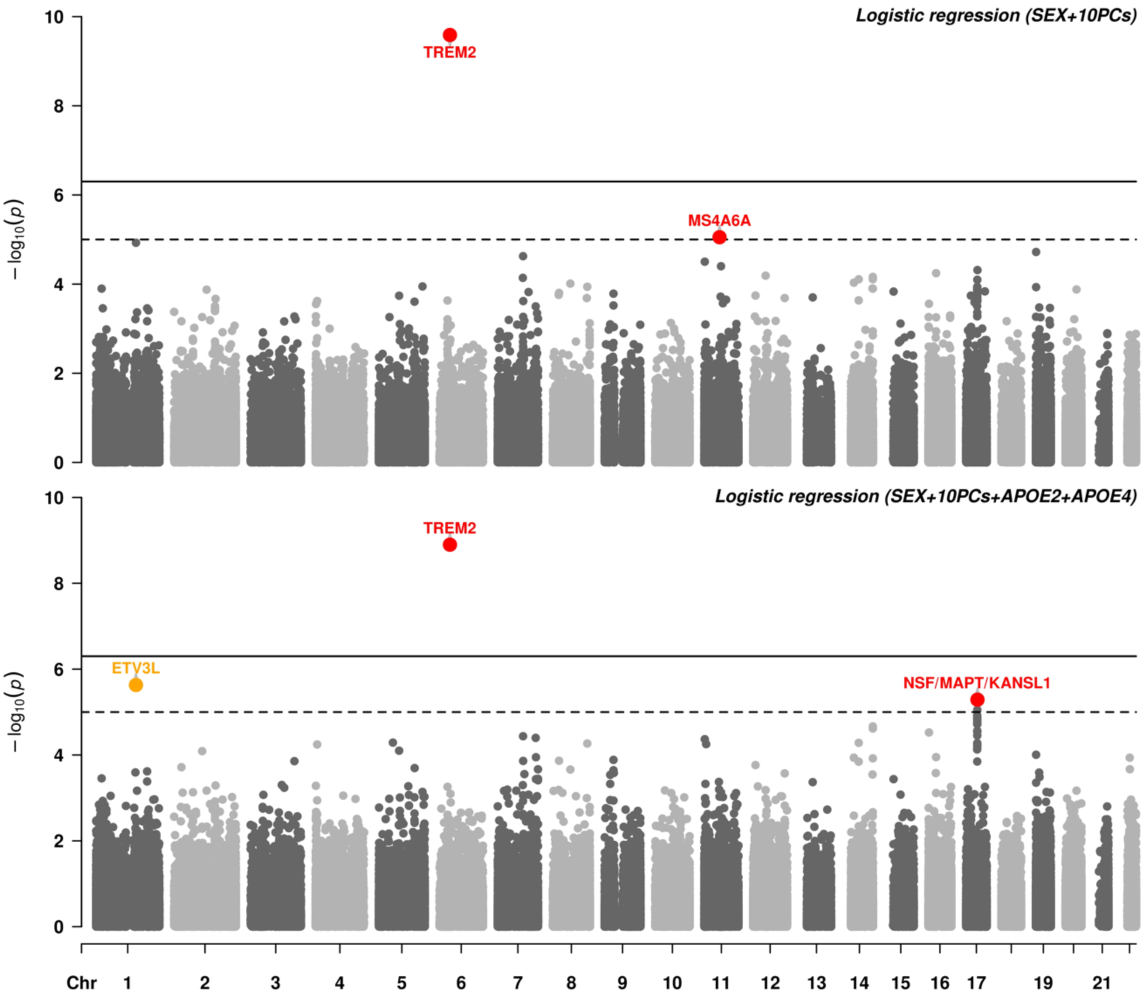


**Figure S4. Manhattan plots for the two model adjustments of the logistic regression not adjusted by age.** The main causal variant on *TREM2* is exome wide significant (solid line, $p < 5\times10^{-7}$). Among suggestive associations (dashed line, $p < 1\times10^{-5}$), (i) known AD associations are in red, (ii) novel associations which replicate ($p < 0.05$) in an independent dataset (cf **Table 3**) are in blue, (iii) likely spurious associations with discordant direction of effect in the replication are in yellow (cf **Table S5**).

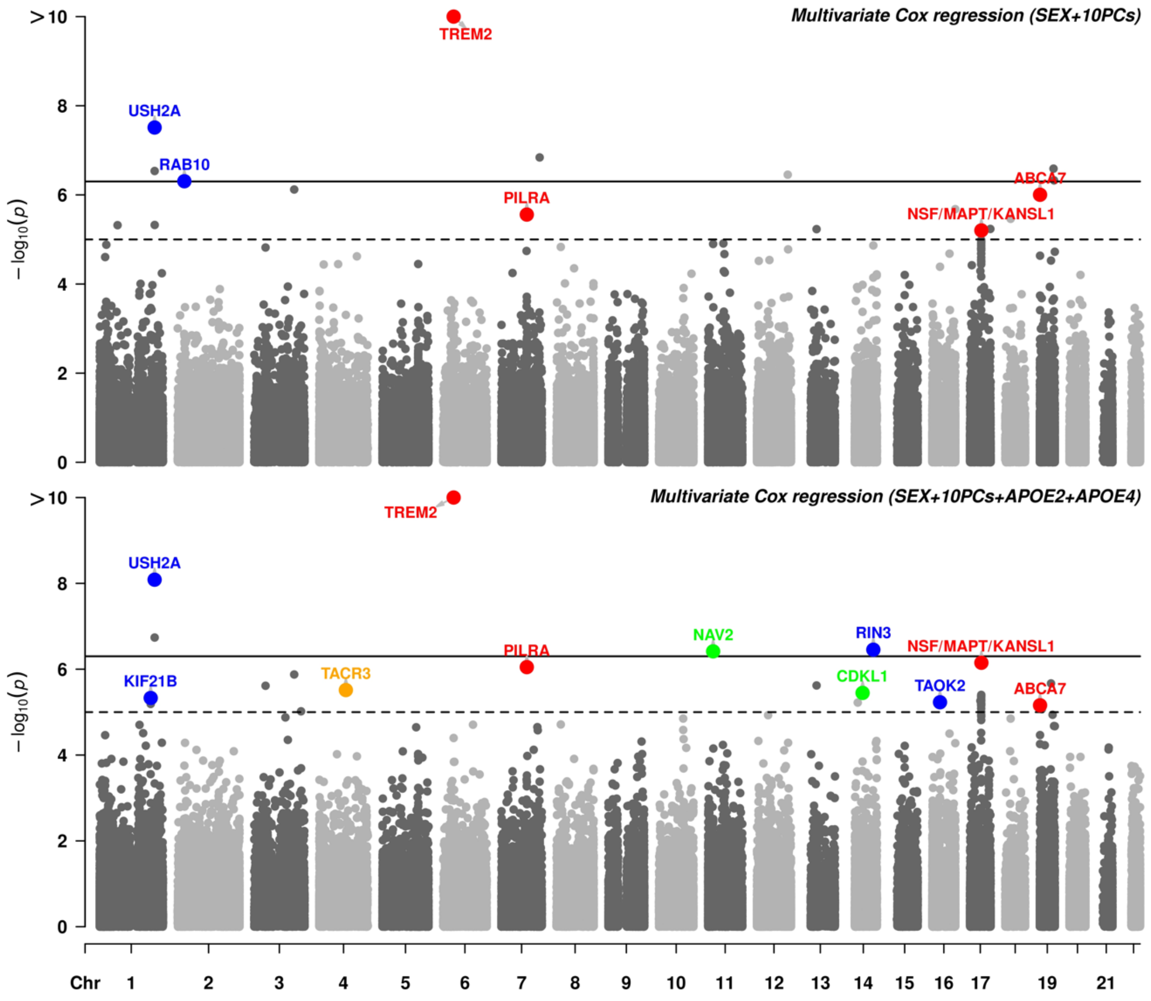


**Figure S5. Manhattan plots for the two model adjustments of the multivariate Cox regression.** The main causal variant on *TREM2* is exome wide significant (solid line, $p < 5\times10^{-7}$). Among suggestive associations (dashed line, $p < 1\times10^{-5}$), (i) known AD associations are in red, (ii) novel associations which replicate ($p < 0.05$) in an independent dataset (cf **Table 3**) are in blue, (iii) associations with concordant direction of effect in the replication which failed to reach nominal significance are in green, (iv) likely spurious associations with discordant direction of effect in the replication are in yellow (cf **Table S5**).

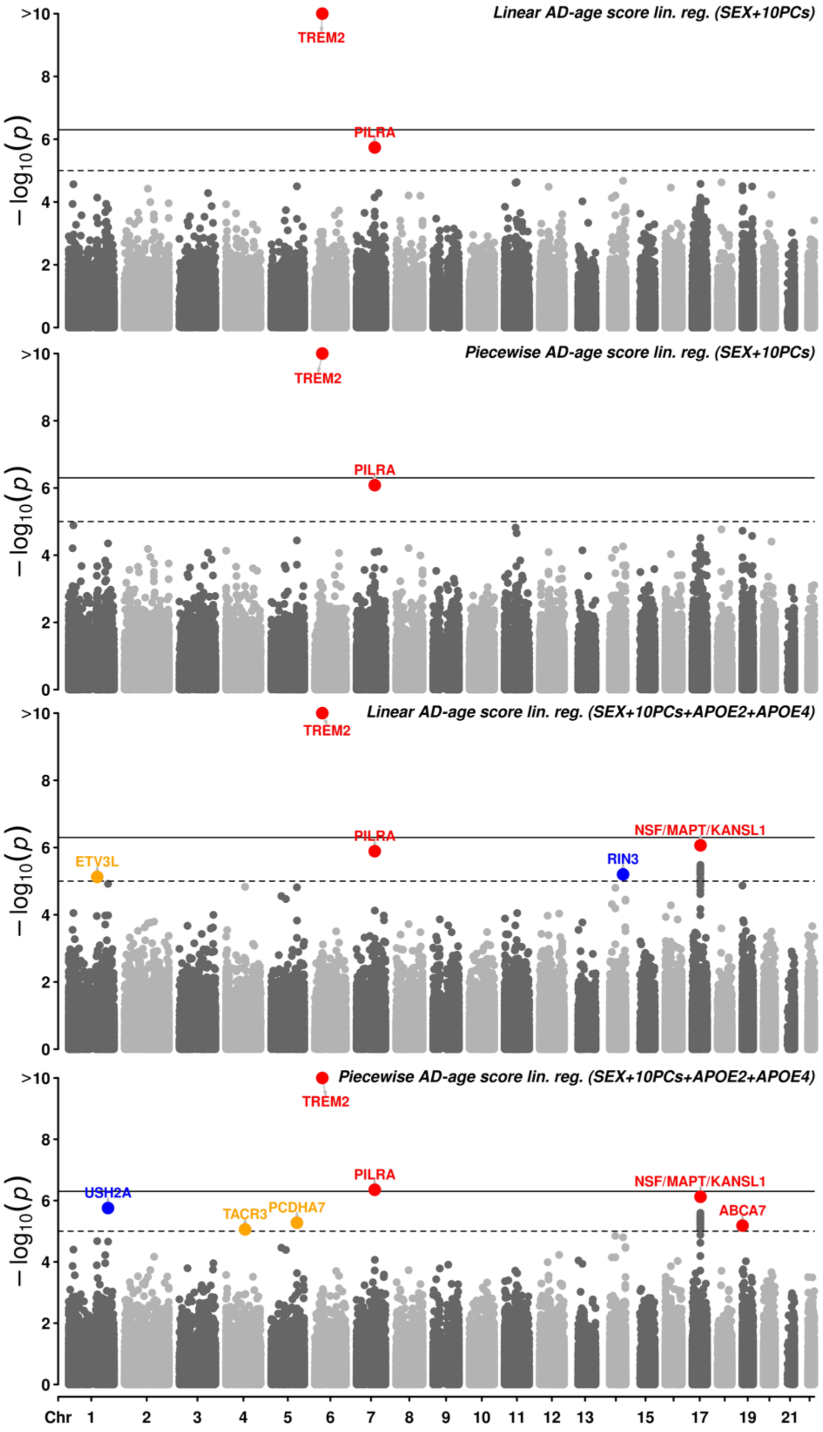


**Figure S6. Manhattan plots for the two model adjustments on the two AD-age scores linear regression.** The main causal variant on *TREM2* is exome wide significant (solid line, $p < 5\times10^{-7}$). Among suggestive associations (dashed line, $p < 1\times10^{-5}$), (i) known AD associations are in red, (ii) novel associations which replicate ($p < 0.05$) in an independent dataset (cf **Table 3**) are in blue, (iii) likely spurious associations with discordant direction of effect in the replication are in yellow (cf **Table S5**). P-values are reported prior to bootstrap based inference, here.

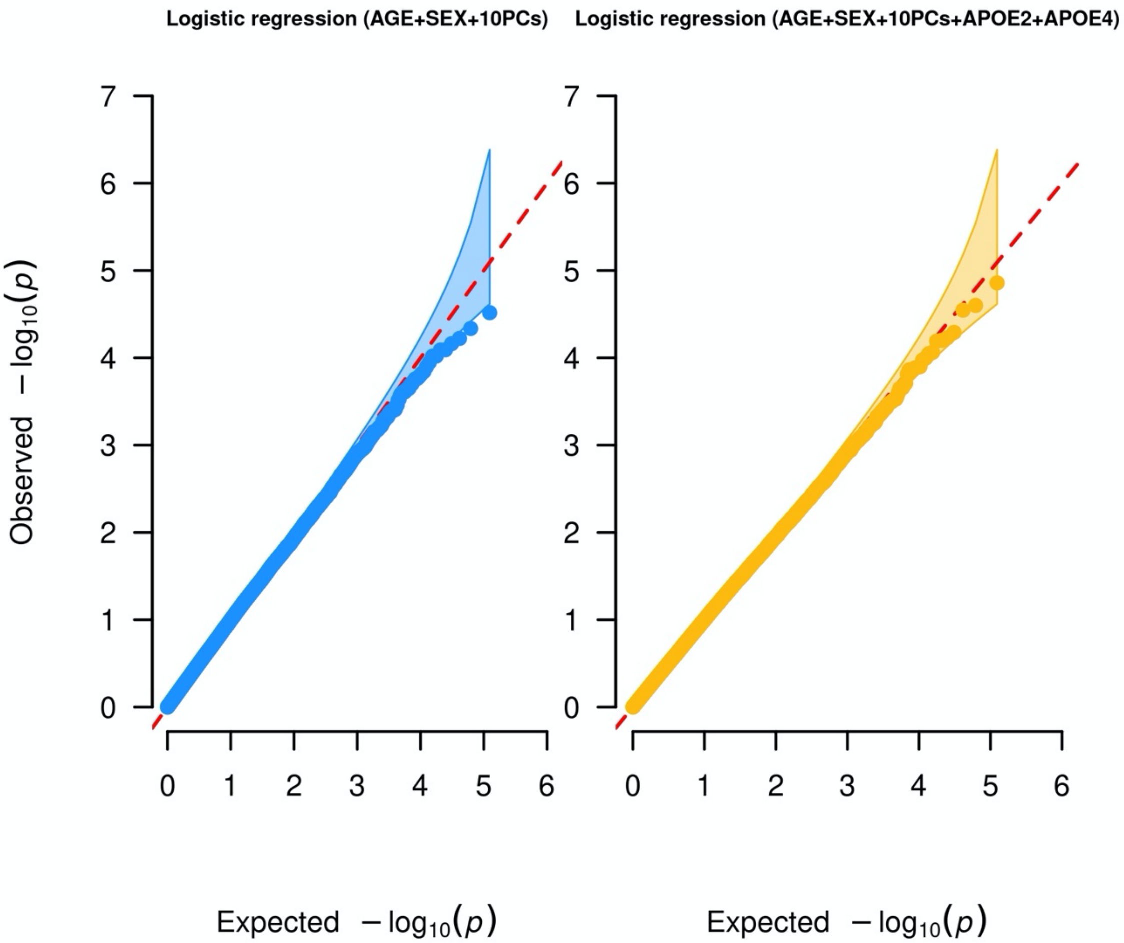


**Figure S7. QQ plots for the logistic regression adjusted by age corresponding to Figure S3.**

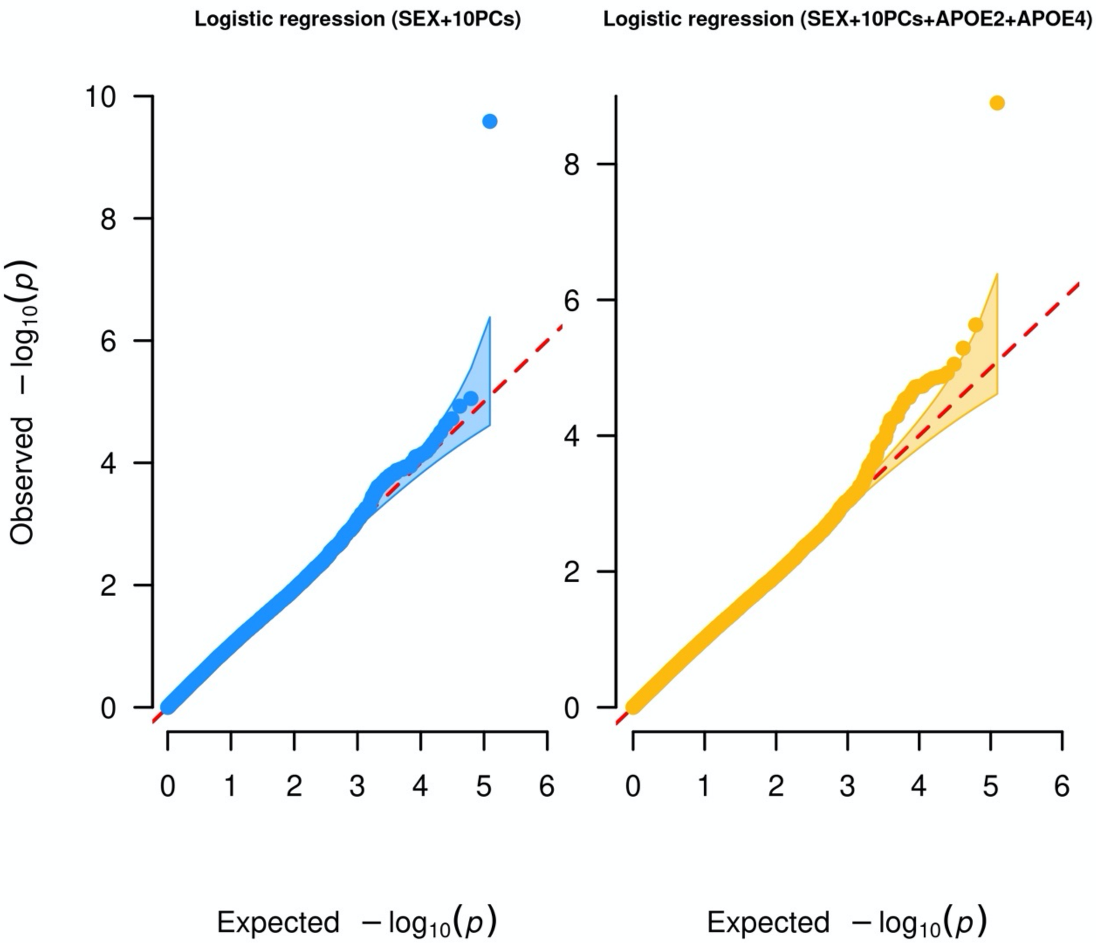


**Figure S8. QQ plots for the standard logistic regression corresponding to Figure S4.**

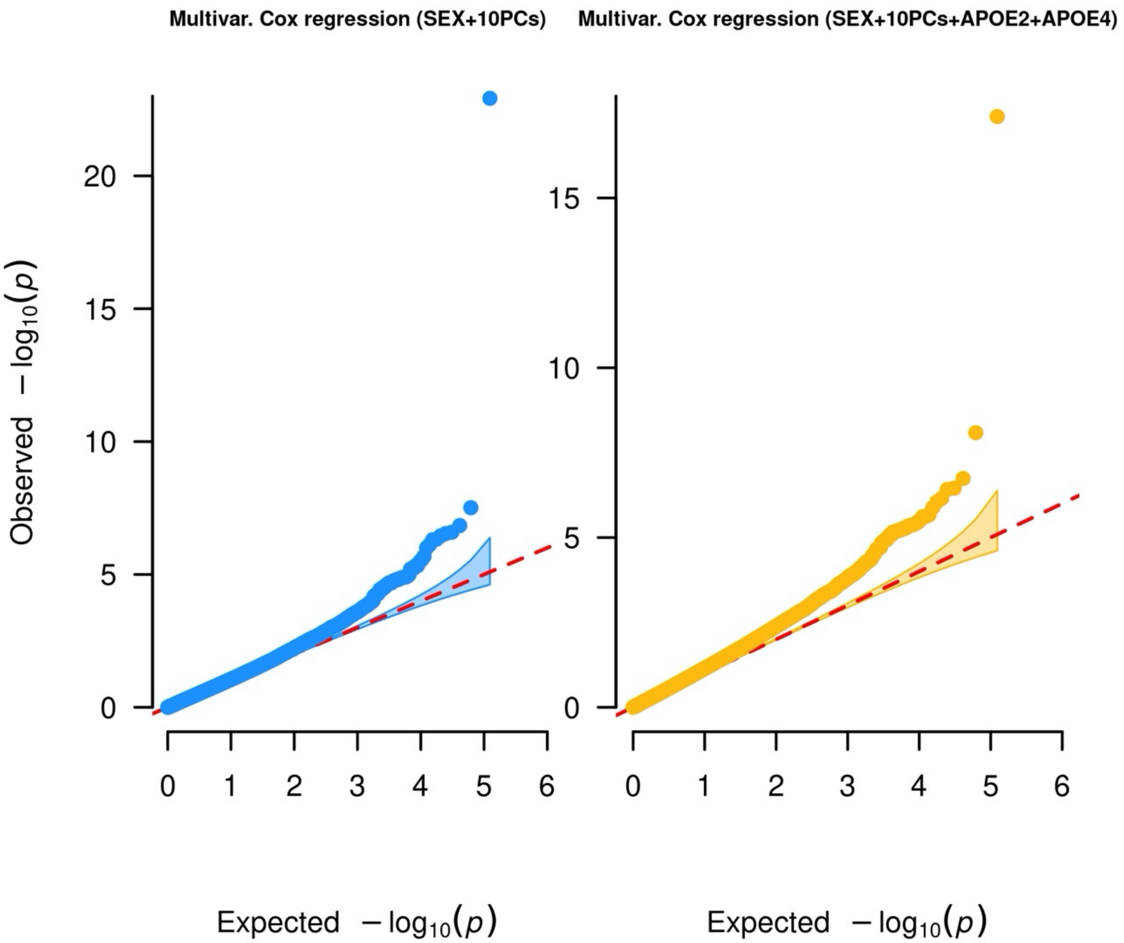


**Figure S9. QQ plots for the multivariate Cox regression corresponding to Figure S5.**

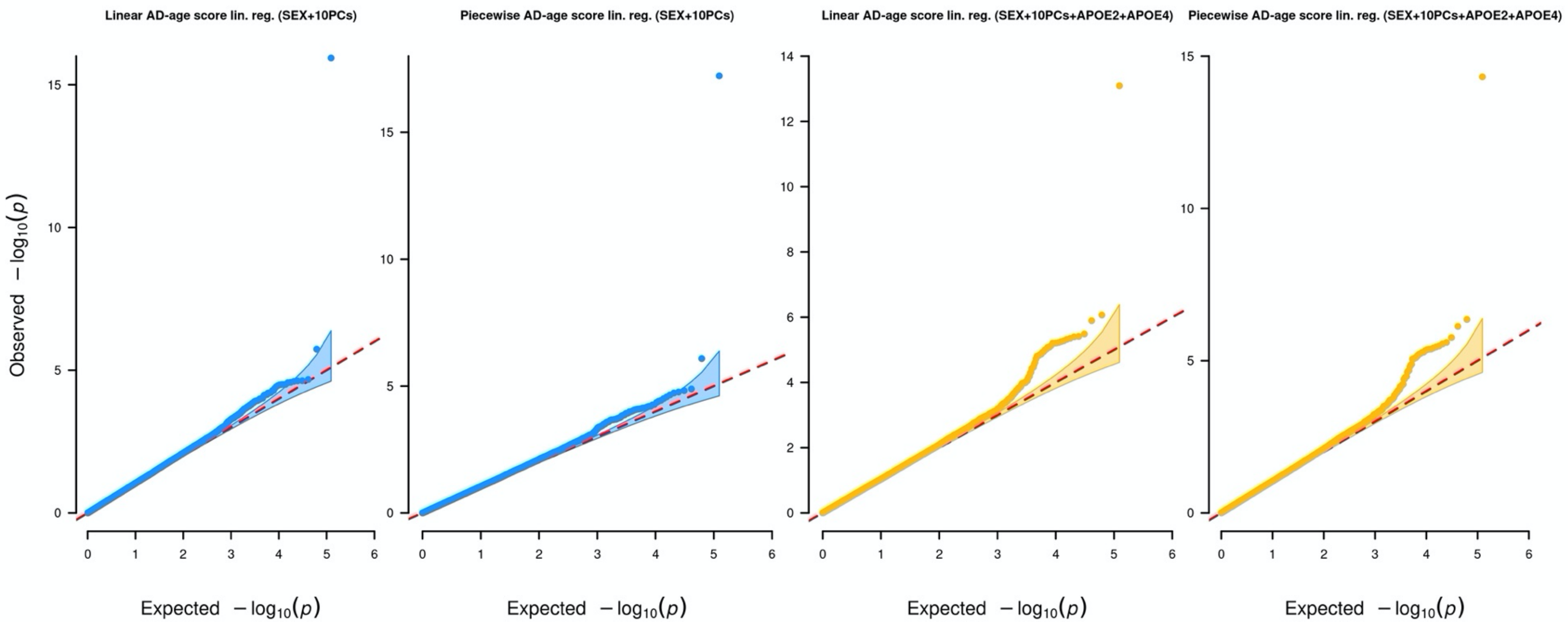


**Figure S10. QQ plots for the linear regression on the AD-age score corresponding to Figure S6.**

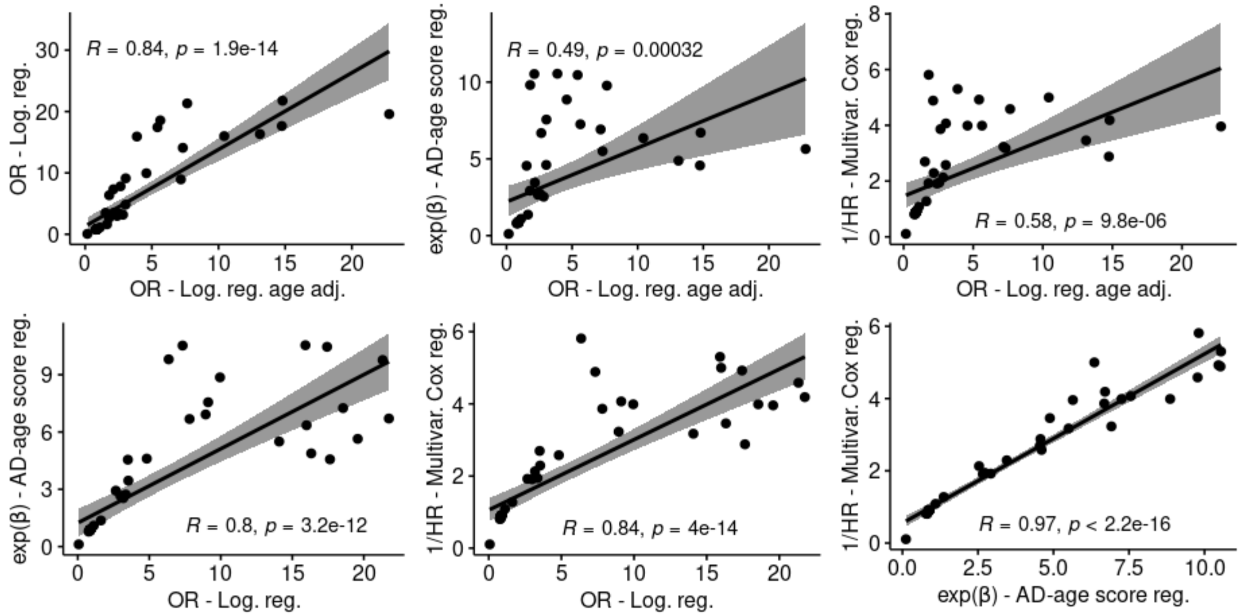


**Figure S11. Comparison between exp(β), OR (odds ratio), 1/HR (hazard ratio) for associations suggestive in any models.**

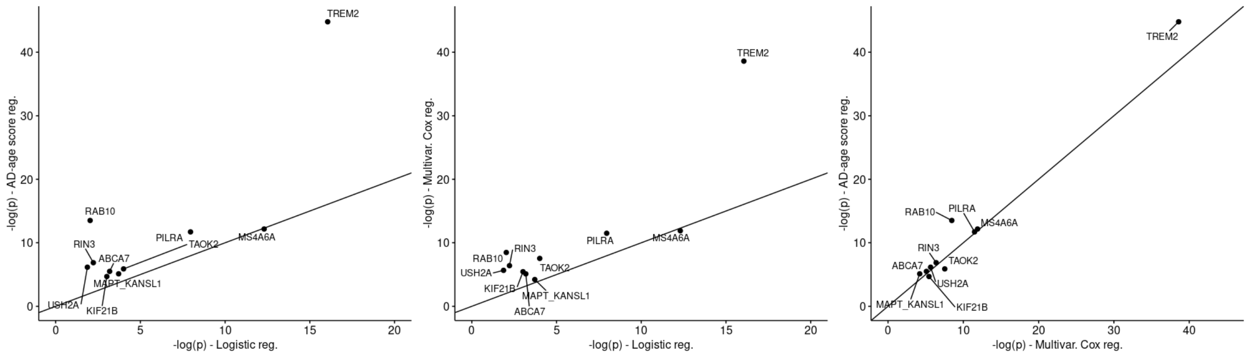


**Figure S12. Comparison between -log(p) between logistic regression, linear regression on the AD-age score and multivariate Cox regression.** Seven out of the ten replicated variants were most significant in the linear regression on the AD-age score, while only two in the Cox regression (on *KIF21B* and *TAOK2*) and one in the logistic regression (on *MS4A6A*). y = x line is represented, variants above this line are most significant in the model in the model on the y-axis.

**Table S1 Demographics per cohort in the discovery sample.**

| Sample | *N* *(% females)* | *Age* μ (σ) | ε3/ε3 (%) | ε3/ε4 (%) | ε4/ε4 (%) | ε2/ε3 (%) | ε2/ε4 (%) | ε2/ε2 (%) |
|---|---|---|---|---|---|---|---|---|
| **Total Discovery** | | | | | | | | |
| Controls | 5075 (59.0) | 85.2 (5.4) | 66.13 | 13.93 | 0.51 | 17.12 | 1.52 | 0.79 |
| AD cases | 6052 (57.8) | 76.3 (8.2) | 47.54 | 39.29 | 4.23 | 6.08 | 2.46 | 0.4 |
| **Discovery** | | | | | | | | |
| ADSP WES $_{controls}$ | 4056 (59.2) | 86.6 (3.7) | 66.62 | 12.25 | 0.35 | 18.34 | 1.48 | 0.96 |
| ADSP WES $_{cases}$ | 4775 (57.8) | 75.9 (8.2) | 50.68 | 37.72 | 2.62 | 6.32 | 2.24 | 0.42 |
| ADSP WGS $_{controls}$ | 677 (60.0) | 78.4 (6.4) | 62.92 | 23.04 | 1.18 | 10.64 | 2.07 | 0.15 |
| ADSP WGS $_{cases}$ | 698 (49.7) | 76.3 (7.7) | 30.09 | 48.57 | 15.04 | 2.72 | 3.44 | 0.14 |
| AMP-AD WGS $_{controls}$ | 342 (54.4) | 81.2 (8.2) | 66.67 | 15.79 | 1.17 | 15.5 | 0.88 | 0 |
| AMP-AD WGS $_{cases}$ | 579 (68.0) | 79.7 (8.0) | 42.66 | 41.11 | 4.49 | 8.12 | 3.11 | 0.52 |

**Table S2. Demographics per cohort in the replication sample.** Each cohort corresponds to a pair of AD study and SNP Array. ADNI_1/O25/OE are part of ADNI. ADM_O/Q are part of ADDNEUROMED. UVM_A/B/C are part of UVM-VU-MSSM. ROSMAP_1B/1T/2 are part of ROSMAP. MAYO_1/2 are part of MAYO. MIRAGE_370/610 are part of MIRAGE. HBTRC_PERL/ILL are part of the HBTRC.

| Sample | *N* (*% females*) | *Age* μ (σ) | ε3/ε3 (%) | ε3/ε4 (%) | ε4/ε4 (%) | ε2/ε3 (%) | ε2/ε4 (%) | ε2/ε2 (%) |
|---|---|---|---|---|---|---|---|---|
| **Total Replication** | | | | | | | | |
| Controls | 10539 (59.4) | 76.7 (8.5) | 60.98 | 22.01 | 2.07 | 12.11 | 2.18 | 0.65 |
| AD cases | 11092 (60.5) | 73.3 (9.3) | 32.83 | 44.37 | 16.21 | 3.69 | 2.79 | 0.1 |
| **Replication** | | | | | | | | |
| ACT $_{controls}$ | 938 (54.6) | 79.4 (5.9) | 62.47 | 21 | 1.28 | 13.22 | 1.71 | 0.32 |
| ACT $_{cases}$ | 289 (64.7) | 81.1 (6.2) | 28.72 | 50.17 | 11.42 | 3.46 | 6.23 | 0 |
| ADC1 $_{controls}$ | 285 (60.0) | 75.8 (11.2) | 55.79 | 28.07 | 2.46 | 11.58 | 1.05 | 1.05 |
| ADC1 $_{cases}$ | 942 (53.9) | 69.0 (11.0) | 24.52 | 46.5 | 24.95 | 2.12 | 1.8 | 0.11 |
| ADC2 $_{controls}$ | 92 (69.6) | 78.0 (9.2) | 58.7 | 16.3 | 3.26 | 15.22 | 5.43 | 1.09 |
| ADC2 $_{cases}$ | 326 (54.3) | 73.0 (7.5) | 25.46 | 43.25 | 25.46 | 2.76 | 3.07 | 0 |
| ADC3 $_{controls}$ | 397 (65.0) | 76.9 (8.7) | 59.95 | 23.17 | 1.76 | 10.83 | 3.02 | 1.26 |
| ADC3 $_{cases}$ | 478 (57.3) | 70.9 (11.2) | 27.41 | 45.61 | 19.67 | 2.93 | 4.18 | 0.21 |
| ADC4 $_{controls}$ | 360 (61.7) | 77.9 (8.5) | 57.5 | 21.94 | 3.06 | 14.72 | 2.22 | 0.56 |
| ADC4 $_{cases}$ | 236 (53.0) | 72.1 (10.3) | 33.9 | 38.14 | 19.07 | 4.66 | 4.24 | 0 |
| ADC5 $_{controls}$ | 410 (65.1) | 79.1 (8.1) | 60 | 21.46 | 2.44 | 13.41 | 2.44 | 0.24 |
| ADC5 $_{cases}$ | 250 (56.8) | 74.4 (9.3) | 32 | 45.6 | 16 | 3.2 | 3.2 | 0 |
| ADC6 $_{controls}$ | 298 (66.4) | 78.2 (8.3) | 62.42 | 26.17 | 1.68 | 8.39 | 1.01 | 0.34 |
| ADC6 $_{cases}$ | 409 (55.0) | 64.5 (12.4) | 38.63 | 39.36 | 16.38 | 3.42 | 1.96 | 0.24 |
| ADC7 $_{controls}$ | 745 (64.0) | 77.5 (7.8) | 59.33 | 26.98 | 1.88 | 8.99 | 2.28 | 0.54 |
| ADC7 $_{cases}$ | 492 (53.3) | 73.0 (7.7) | 32.32 | 44.72 | 14.43 | 5.28 | 3.25 | 0 |
| ADMO $_{controls}$ | 113 (55.8) | 77.6 (7.2) | 70.8 | 16.81 | 0.88 | 7.96 | 2.65 | 0.88 |
| ADMO $_{cases}$ | 125 (67.2) | 74.5 (6.8) | 44 | 40 | 8.8 | 4 | 3.2 | 0 |
| ADMQ $_{controls}$ | 71 (59.2) | 74.5 (5.1) | 57.75 | 26.76 | 2.82 | 11.27 | 1.41 | 0 |
| ADMQ $_{cases}$ | 127 (62.2) | 71.6 (6.4) | 38.58 | 42.52 | 14.17 | 3.94 | 0.79 | 0 |
| ADNI1 $_{controls}$ | 72 (40.3) | 78.4 (6.1) | 61.11 | 20.83 | 5.56 | 11.11 | 0 | 1.39 |
| ADNI1 $_{cases}$ | 235 (42.6) | 75.2 (7.6) | 29.79 | 46.38 | 18.3 | 2.55 | 2.98 | 0 |
| ADNI_DOD $_{controls}$ | 79 (0.0) | 70.2 (5.3) | 56.96 | 20.25 | 2.53 | 18.99 | 1.27 | 0 |
| ADNI_O25 $_{controls}$ | 18 (38.9) | 80.0 (8.6) | 72.22 | 22.22 | 0 | 5.56 | 0 | 0 |
| ADNI_O25 $_{cases}$ | 27 (40.7) | 70.1 (11.1) | 33.33 | 51.85 | 14.81 | 0 | 0 | 0 |

| Sample | *N* (*% females*) | *Age* μ (σ) | ε3/ε3 (%) | ε3/ε4 (%) | ε4/ε4 (%) | ε2/ε3 (%) | ε2/ε4 (%) | ε2/ε2 (%) |
|---|---|---|---|---|---|---|---|---|
| ADNI_OE controls | 97 (57.7) | 76.1 (5.9) | 56.7 | 27.84 | 1.03 | 11.34 | 3.09 | 0 |
| ADNI_OE cases | 118 (43.2) | 73.2 (7.4) | 26.27 | 46.61 | 19.49 | 5.08 | 2.54 | 0 |
| CIDR controls | 95 (62.1) | 77.1 (8.2) | 69.47 | 14.74 | 3.16 | 11.58 | 1.05 | 0 |
| CIDR cases | 50 (66.0) | 77.1 (6.8) | 56 | 30 | 6 | 6 | 2 | 0 |
| GenADA controls | 769 (64.2) | 73.5 (7.9) | 63.46 | 20.16 | 1.56 | 11.7 | 2.6 | 0.52 |
| GenADA cases | 779 (57.6) | 72.6 (8.6) | 32.86 | 46.34 | 13.35 | 4.36 | 3.08 | 0 |
| HBTRC_A controls | 81 (24.7) | 65.3 (9.6) | 59.26 | 28.4 | 3.7 | 6.17 | 2.47 | 0 |
| HBTRC_A cases | 181 (59.7) | 70.2 (8.5) | 39.23 | 45.3 | 7.18 | 3.87 | 4.42 | 0 |
| HBTRC_B controls | 52 (21.2) | 61.9 (7.8) | 48.08 | 30.77 | 11.54 | 7.69 | 1.92 | 0 |
| HBTRC_B cases | 101 (49.5) | 70.4 (8.7) | 46.53 | 34.65 | 6.93 | 7.92 | 3.96 | 0 |
| LOAD controls | 445 (60.7) | 75.9 (8.3) | 57.3 | 25.84 | 1.8 | 11.24 | 3.37 | 0.45 |
| LOAD cases | 1422 (65.9) | 73.3 (7.2) | 21.94 | 51.41 | 22.22 | 2.04 | 2.25 | 0.14 |
| MARS controls | 148 (77.0) | 82.5 (7.8) | 62.84 | 18.92 | 0.68 | 12.84 | 3.38 | 1.35 |
| MARS cases | 70 (80.0) | 85.1 (7.6) | 58.57 | 27.14 | 1.43 | 11.43 | 1.43 | 0 |
| MAYO controls | 1069 (50.8) | 73.1 (4.5) | 58.19 | 24.23 | 1.87 | 11.79 | 3.27 | 0.65 |
| MAYO cases | 495 (61.6) | 74.0 (5.0) | 25.25 | 47.68 | 22.63 | 1.82 | 2.63 | 0 |
| MAYO2 controls | 130 (46.2) | 77.5 (8.3) | 64.62 | 16.92 | 0.77 | 16.15 | 0.77 | 0.77 |
| MAYO2 cases | 4 (50.0) | 71.2 (14.3) | 75 | 25 | 0 | 0 | 0 | 0 |
| MIRAGE_370 controls | 146 (64.4) | 71.6 (7.0) | 47.26 | 36.3 | 5.48 | 8.9 | 2.05 | 0 |
| MIRAGE_370 cases | 79 (49.4) | 65.7 (9.3) | 36.71 | 37.97 | 21.52 | 3.8 | 0 | 0 |
| MIRAGE_610 controls | 534 (56.0) | 71.9 (7.1) | 54.87 | 29.21 | 6.18 | 8.05 | 1.5 | 0.19 |
| MIRAGE_610 cases | 233 (63.1) | 67.9 (9.6) | 33.05 | 41.63 | 18.03 | 3.43 | 3.86 | 0 |
| OHSU controls | 217 (53.5) | 86.9 (7.5) | 59.45 | 18.43 | 1.38 | 17.97 | 2.76 | 0 |
| OHSU cases | 68 (57.4) | 85.4 (9.2) | 51.47 | 36.76 | 2.94 | 7.35 | 1.47 | 0 |
| ROSMAP2 controls | 143 (75.5) | 85.3 (7.2) | 65.03 | 16.08 | 1.4 | 15.38 | 1.4 | 0.7 |
| ROSMAP2 cases | 74 (79.7) | 85.5 (5.8) | 67.57 | 21.62 | 4.05 | 5.41 | 1.35 | 0 |
| ROSMAP1B controls | 484 (73.3) | 87.1 (7.2) | 64.88 | 14.26 | 1.24 | 18.18 | 1.03 | 0.41 |
| ROSMAP1B cases | 418 (72.5) | 83.7 (6.9) | 56.7 | 28.47 | 2.39 | 10.53 | 1.67 | 0.24 |
| ROSMAP1T controls | 34 (52.9) | 83.0 (7.1) | 70.59 | 14.71 | 0 | 14.71 | 0 | 0 |
| ROSMAP1T cases | 116 (63.8) | 84.8 (5.4) | 50 | 37.07 | 2.59 | 6.9 | 3.45 | 0 |
| TGEN controls | 398 (48.0) | 80.2 (8.9) | 62.81 | 17.84 | 1.76 | 12.06 | 2.26 | 3.27 |
| TGEN cases | 752 (65.8) | 72.6 (8.0) | 30.59 | 45.21 | 17.55 | 3.06 | 3.32 | 0.27 |
| UPITT controls | 879 (63.0) | 75.5 (6.1) | 65.98 | 16.38 | 1.14 | 13.88 | 2.05 | 0.57 |
| UPITT cases | 1301 (63.3) | 73.3 (6.6) | 38.28 | 45.35 | 9.76 | 3.54 | 2.84 | 0.23 |
| UVM_A controls | 453 (68.2) | 73.0 (6.7) | 60.71 | 22.74 | 1.55 | 12.58 | 1.77 | 0.66 |

| Sample | *N* (*% females*) | *Age* μ (σ) | ε3/ε3 (%) | ε3/ε4 (%) | ε4/ε4 (%) | ε2/ε3 (%) | ε2/ε4 (%) | ε2/ε2 (%) |
|---|---|---|---|---|---|---|---|---|
| UVM_A cases | 82 (62.2) | 73.3 (6.6) | 10.98 | 45.12 | 43.9 | 0 | 0 | 0 |
| UVM_B controls | 235 (60.4) | 74.6 (9.2) | 67.66 | 18.72 | 0.85 | 9.79 | 2.13 | 0.85 |
| UVM_B cases | 240 (70.0) | 77.1 (9.1) | 46.67 | 34.58 | 12.08 | 4.58 | 2.08 | 0 |
| UVM_C controls | 109 (47.7) | 75.8 (9.1) | 67.89 | 18.35 | 0.92 | 9.17 | 1.83 | 1.83 |
| UVM_C cases | 278 (62.9) | 72.2 (7.5) | 32.01 | 46.04 | 17.99 | 2.16 | 1.8 | 0 |
| WASHU controls | 143 (61.5) | 76.9 (8.5) | 62.94 | 20.98 | 4.2 | 9.79 | 1.4 | 0.7 |
| WASHU cases | 295 (56.9) | 75.3 (9.9) | 39.32 | 42.37 | 8.14 | 6.44 | 3.73 | 0 |

**Table S3. Sample sizes, minor allele frequency and imputation quality for the suggestive variants in the discovery.** MAF: Minor allele frequency; R-square (Rsq): Imputation quality.

| Gene(s) | RS id | SNP (hg19) | Discovery | | Replication | | |
|---|---|---|---|---|---|---|---|
| | | | N | MAF | N | MAF | Rsq |
| ***ANKRD13C*** | rs777614616 | 1:70801760:A:C | 11096 | 0.00054 | 6495 | 0.00064 | 0.83 |
| ***ETV3L*** | rs16838078 | 1:157069261:A:C | 9746 | 0.02427 | 21631 | 0.02724 | 0.96 |
| ***KIF21B*** | rs2297911 | 1:200959302:G:A | 11006 | 0.17391 | 21631 | 0.1769 | 1 |
| ***USH2A*** | rs111033333 | 1:216270469:G:A | 11126 | 0.00085 | 19544 | 0.00132 | 0.81 |
| ***RAB10*** | rs149622307 | 2:26332640:T:C | 11057 | 0.00045 | 9833 | 0.00076 | 0.85 |
| ***ZNF197*** | rs140904465 | 3:44685500:T:G | 11126 | 0.00094 | 20403 | 0.00132 | 0.86 |
| ***VEPH1*** | rs138491831 | 3:157178046:G:A | 11126 | 0.00054 | 16219 | 0.00101 | 0.86 |
| ***CHRD*** | rs3749228 | 3:184099342:G:C | 10793 | 0.0504 | 21631 | 0.05688 | 0.96 |
| ***TACR3*** | rs144292455 | 4:104577415:C:T | 11105 | 0.00081 | 17657 | 0.001 | 0.86 |
| ***PCDHA7*** | rs143048298 | 5:140215922:A:G | 11122 | 0.00067 | 15845 | 0.00074 | 0.92 |
| ***TREM2*** | rs75932628 | 6:41129252:C:T | 11076 | 0.00591 | 21176 | 0.00606 | 0.93 |
| ***PILRA*** | rs2405442 | 7:99971313:T:C | 11022 | 0.29836 | 21631 | 0.30567 | 0.94 |
| ***GIMAP2*** | rs778956614 | 7:150389837:TC:T | 11126 | 0.00045 | 17015 | 0.00081 | 0.87 |
| ***NAV2*** | rs11828836 | 11:19735325:C:A | 11105 | 0.00045 | 9235 | 0.00045 | 0.9 |
| ***MS4A6A*** | rs12453 | 11:59945745:T:C | 11114 | 0.3941 | 21481 | 0.39015 | 0.99 |
| ***COQ5*** | rs74578594 | 12:120947933:G:C | 11126 | 0.00049 | 13736 | 0.00075 | 0.94 |
| ***DGKH*** | rs147914294 | 13:42761244:C:A | 11126 | 0.0031 | 21486 | 0.00322 | 0.94 |
| ***HEATR5A*** | rs28396248 | 14:31790886:T:G | 10673 | 0.04572 | 21631 | 0.06432 | 0.99 |
| ***CDKL1*** | rs61981931 | 14:50856882:C:T | 11110 | 0.04977 | 21631 | 0.04392 | 0.81 |

| Gene(s) | RS id | SNP (hg19) | Discovery | | Replication | | |
|---|---|---|---|---|---|---|---|
| | | | N | MAF | N | MAF | Rsq |
| ***RIN3*** | rs150221413 | 14:93022240:G:T | 11020 | 0.00082 | 17652 | 0.00131 | 0.8 |
| ***TAOK2*** | rs4077410 | 16:29998200:A:G | 11063 | 0.47966 | 21631 | 0.48195 | 0.94 |
| ***MC1R*** | rs34158934 | 16:89985950:C:T | 11024 | 0.00082 | 13553 | 0.00062 | 0.77 |
| ***AOC2*** | rs201046755 | 17:40997372:G:A | 10206 | 0.00054 | 14425 | 0.00058 | 0.85 |
| ***NSF/MAPT/KANSL1*** | rs199533 | 17:44828931:G:A | 11094 | 0.20367 | 21631 | 0.19931 | 0.99 |
| ***RFNG*** | rs112510774 | 17:80008395:G:A | 11073 | 0.00117 | 21070 | 0.00204 | 0.77 |
| ***C18orf8*** | rs112277818 | 18:21083647:C:A | 11110 | 0.00302 | 21015 | 0.0031 | 0.86 |
| ***ABCA7*** | rs547447016 | 19:1047507:AGGAGCAG:A | 11006 | 0.00313 | 18356 | 0.00311 | 0.88 |
| ***CAMSAP3*** | rs144062687 | 19:7682224:G:A | 11087 | 0.00198 | 19642 | 0.00285 | 0.69 |
| ***SRRM5*** | rs200590643 | 19:44116708:A:G | 10127 | 0.00049 | 14411 | 0.00109 | 0.66 |
| ***ZNF765*** | rs140062377 | 19:53912054:T:C | 11122 | 0.00216 | 18673 | 0.00194 | 0.6 |
| ***SSC5D*** | rs776353690 | 19:56005235:A:G | 11087 | 0.0005 | 18005 | 0.00089 | 0.85 |

**Table S4. Lambda medians for each main model and model adjustments.**

| Model | Lambda median |
|---|---|
| Logistic regression (AGE+SEX+10PCs) | 1.02 |
| Logistic regression (AGE+SEX+10PCs+APOE2+APOE4) | 1.01 |
| Logistic regression (SEX+10PCs) | 1.06 |
| Logistic regression (SEX+10PCs+APOE2+APOE4) | 1.05 |
| Multivariate Cox regression (SEX+10PCs) | 1.09 |
| Multivariate Cox regression (SEX+10PCs+APOE2+APOE4) | 1.19 |
| Linear AD-age score lin. reg. (SEX+10PCs) | 1.08 |
| Piecewise AD-age score lin. reg. (SEX+10PCs) | 1.07 |
| Linear AD-age score lin. reg. (SEX+10PCs+APOE2+APOE4) | 1.06 |
| Piecewise AD-age score lin. reg. (SEX+10PCs+APOE2+APOE4) | 1.06 |

**Table S5. All suggestive association results in the discovery.** Effect corresponds to OR (odds ratio) for logistic regression on AD status not adjusted by age (LogReg), exp(β) for linear regression on AD-age score (LinReg), and 1/HR (hazard ratio) for multivariate Cox regression on age-at-onset (CoxReg). Correlation between these measures is high for suggestive associations as shown on **Figure S11**. P: p-value. m: model subversion. Subversion codes are: (1) adjusted for sex and 10 first principal components of population structure and (2) additionally adjusted for *APOE* ε2/ε4 alleles. Two types of weighted AD-age score were used with (A) corresponding to a linear effect of age between 60 and 100 and (B) accounting for the changes in AD prevalence slope in this age range. P-values are reported here before bootstrapping for LinReg.

| SNP (hg19) | Discovery | | | | | | | | | Replication | | | | | | | | |
|---|---|---|---|---|---|---|---|---|---|---|---|---|---|---|---|---|---|---|
| | LogReg | | | LinReg | | | CoxReg | | | LogReg | | | LinReg | | | CoxReg | | |
| | OR | P | m | exp(β) | P | m | 1/HR | P | m | OR | P | m | exp(β) | P | m | 1/HR | P | m |
| 1:70801760:A:C | 9.95 | 0.03 | 1 | 8.86 | $2.10^{-4}$ | B1 | 4.00 | $5.10^{-6}$ | 1 | 0.58 | 40.46 | 2 | 0.63 | 0.33 | A2 | 0.80 | 0.65 | 2 |
| 1:157069261:A:C | 1.62 | $2.10^{-6}$ | 2 | 1.37 | $7.10^{-6}$ | A2 | 1.28 | $2.10^{-5}$ | 2 | 0.92 | 0.14 | 1 | 0.92 | 0.11 | A1 | 0.94 | 10.14 | 1 |
| 1:200959302:G:A | 0.87 | $2.10^{-4}$ | 2 | 0.88 | $6.10^{-5}$ | B2 | 0.89 | $5.10^{-6}$ | 2 | 0.96 | 0.13 | 1 | 0.95 | 0.04 | B1 | 0.96 | 0.02 | 2 |
| 1:216270469:G:A | 9.12 | $4.10^{-3}$ | 2 | 7.56 | $2.10^{-6}$ | B2 | 4.07 | $8.10^{-9}$ | 2 | 1.58 | 0.14 | 1 | 1.70 | 0.04 | A1 | 1.33 | 0.12 | 1 |
| 2:26332640:T:C | 17.4 | 0.06 | 1 | 10.5 | $3.10^{-4}$ | B1 | 4.92 | $5.10^{-7}$ | 1 | 4.50 | 0.05 | 1 | 5.03 | $3.10^{-3}$ | B1 | 2.69 | $6.10^{-4}$ | 1 |
| 3:44685500:T:G | 17.64 | $5.10^{-3}$ | 2 | 4.57 | $2.10^{-4}$ | B2 | 2.88 | $2.10^{-6}$ | 2 | 0.74 | 0.28 | 1 | 0.89 | 0.63 | B1 | 0.86 | 0.46 | 2 |
| 3:157178046:G:A | 21.76 | 0.04 | 1 | 6.71 | $5.10^{-5}$ | A1 | 4.19 | $8.10^{-7}$ | 1 | 1.16 | 0.72 | 2 | 0.91 | 0.79 | B1 | 1.24 | 0.43 | 2 |
| 3:184099342:G:C | 0.83 | $2.10^{-3}$ | 1 | 0.81 | $2.10^{-4}$ | B2 | 0.82 | $1.10^{-5}$ | 2 | 0.95 | 0.29 | 2 | 0.98 | 0.51 | B2 | 0.97 | 0.31 | 1 |
| 4:104577415:C:T | 8.94 | $4.10^{-3}$ | 2 | 6.92 | $9.10^{-6}$ | B2 | 3.23 | $3.10^{-6}$ | 2 | 0.72 | 0.34 | 1 | 0.81 | 0.54 | B1 | 1.29 | 0.32 | 2 |
| 5:140215922:A:G | 0.069 | $1.10^{-3}$ | 2 | 0.11 | $5.10^{-6}$ | B2 | 0.107 | $2.10^{-3}$ | 2 | 1.14 | 0.76 | 2 | 1.24 | 0.5 | A2 | 0.93 | 0.79 | 2 |
| 6:41129252:C:T | 4.83 | $3.10^{-10}$ | 1 | 4.60 | $6.10^{-18}$ | B1 | 2.58 | $1.10^{-23}$ | 1 | 2.32 | $2.10^{-9}$ | 1 | 2.46 | $1.10^{-15}$ | A1 | 1.95 | $2.10^{-18}$ | 2 |
| 7:99971313:T:C | 0.88 | $2.10^{-5}$ | 1 | 0.87 | $4.10^{-7}$ | B2 | 0.90 | $9.10^{-7}$ | 2 | 0.92 | $6.10^{-5}$ | 1 | 0.90 | $2.10^{-6}$ | B1 | 0.93 | $5.10^{-7}$ | 1 |
| 7:150389837:TC:T | 15.9 | 0.07 | 1 | 10.5 | $3.10^{-4}$ | B1 | 5.3 | $1.10^{-7}$ | 1 | 1.40 | 0.42 | 2 | 0.75 | 0.42 | A1 | 0.83 | 0.46 | 1 |
| 11:59945745:T:C | 0.88 | $9.10^{-6}$ | 1 | 0.89 | $1.10^{-5}$ | B1 | 0.92 | $1.10^{-5}$ | 1 | 0.89 | $1.10^{-8}$ | 1 | 0.89 | $7.10^{-9}$ | B1 | 0.93 | $2.10^{-8}$ | 1 |
| 12:120947933:G:C | 6.36 | 0.02 | 1 | 9.81 | $3.10^{-4}$ | B1 | 5.81 | $4.10^{-7}$ | 1 | 1.12 | 0.83 | 2 | 1.83 | 0.14 | A2 | 1.61 | 0.13 | 1 |
| 13:42761244:C:A | 2.98 | $2.10^{-4}$ | 1 | 2.66 | $7.10^{-5}$ | B1 | 1.91 | $2.10^{-6}$ | 2 | 1.13 | 0.54 | 2 | 1.13 | 0.39 | A2 | 0.99 | 0.96 | 2 |

| SNP (hg19) | **Discovery** | | | | | | | | | **Replication** | | | | | | | | |
|---|---|---|---|---|---|---|---|---|---|---|---|---|---|---|---|---|---|---|
| | **LogReg** | | | **LinReg** | | | **CoxReg** | | | **LogReg** | | | **LinReg** | | | **CoxReg** | | |
| | **OR** | **P** | **m** | **exp(β)** | **P** | **m** | **1/HR** | **P** | **m** | **OR** | **P** | **m** | **exp(β)** | **P** | **m** | **1/HR** | **P** | **m** |
| 14:31790886:T:G | 0.77 | $9.10^{-5}$ | 1 | 0.82 | $5.10^{-5}$ | A2 | 0.81 | $6.10^{-6}$ | 2 | 1.01 | 0.88 | 2 | 0.99 | 0.82 | A1 | 0.98 | 0.40 | 2 |
| 14:50856882:C:T | 0.77 | $5.10^{-5}$ | 2 | 0.78 | $1.10^{-5}$ | B2 | 0.82 | $4.10^{-6}$ | 2 | 0.94 | 0.20 | 1 | 0.94 | 0.13 | A1 | 0.95 | 0.10 | 1 |
| 14:93022240:G:T | 16.3 | $7.10^{-3}$ | 2 | 4.88 | $6.10^{-6}$ | A2 | 3.46 | $4.10^{-7}$ | 2 | 1.95 | 0.04 | 2 | 1.69 | 0.05 | B2 | 1.59 | 0.01 | 2 |
| 16:29998200:A:G | 1.12 | $6.10^{-5}$ | 1 | 1.09 | $3.10^{-5}$ | A1 | 1.09 | $6.10^{-6}$ | 2 | 1.04 | 0.07 | 2 | 1.05 | $3.10^{-3}$ | A2 | 1.05 | $4.10^{-4}$ | 2 |
| 16:89985950:C:T | 14.1 | 0.01 | 1 | 5.50 | $4.10^{-4}$ | B1 | 3.17 | $2.10^{-6}$ | 1 | 1.28 | 0.67 | 1 | 1.38 | 0.49 | A2 | 1.32 | 0.46 | 2 |
| 17:40997372:G:A | 19.6 | 0.05 | 1 | 5.64 | $5.10^{-4}$ | A1 | 3.96 | $5.10^{-6}$ | 2 | 0.69 | 0.53 | 1 | 0.65 | 0.42 | A1 | 0.73 | 0.49 | 1 |
| 17:44828931:G:A | 0.85 | $5.10^{-6}$ | 2 | 0.86 | $8.10^{-7}$ | B2 | 0.89 | $7.10^{-7}$ | 2 | 0.97 | 0.02 | 2 | 0.97 | 0.017 | A2 | 0.98 | 0.018 | 2 |
| 17:80008395:G:A | 3.50 | 0.01 | 1 | 4.55 | $2.10^{-4}$ | B1 | 2.70 | $6.10^{-6}$ | 1 | 0.68 | 0.08 | 1 | 0.87 | 0.46 | A1 | 0.95 | 0.75 | 2 |
| 18:21083647:C:A | 2.66 | $7.10^{-4}$ | 1 | 2.93 | $2.10^{-5}$ | B1 | 1.92 | $3.10^{-6}$ | 1 | 0.64 | 0.02 | 1 | 0.77 | 0.15 | B1 | 0.86 | 0.32 | 1 |
| 19:1047507:AGGAGCAG:A | 3.36 | $1.10^{-4}$ | 2 | 2.73 | $7.10^{-6}$ | B2 | 1.94 | $1.10^{-6}$ | 1 | 1.36 | 0.12 | 1 | 1.33 | 0.14 | B1 | 1.22 | 0.13 | 2 |
| 19:7682224:G:A | 3.18 | $2.10^{-3}$ | 1 | 2.54 | $2.10^{-4}$ | A1 | 2.13 | $9.10^{-6}$ | 2 | 1.18 | 0.47 | 2 | 1.14 | 0.50 | B2 | 1.16 | 0.32 | 2 |
| 19:44116708:A:G | 7.33 | 0.06 | 1 | 10.53 | $3.10^{-4}$ | B1 | 4.89 | $2.10^{-6}$ | 2 | 1.09 | 0.81 | 1 | 0.86 | 0.61 | A2 | 0.98 | 0.93 | 2 |
| 19:53912054:T:C | 3.54 | $7.10^{-4}$ | 1 | 3.45 | $3.10^{-5}$ | B1 | 2.29 | $3.10^{-7}$ | 1 | 0.81 | 0.47 | 1 | 0.81 | 0.40 | A1 | 1.22 | 0.35 | 2 |
| 19:56005235:A:G | 21.3 | 0.04 | 1 | 9.78 | $2.10^{-4}$ | B1 | 4.58 | $5.10^{-7}$ | 1 | 1.91 | 0.12 | 1 | 1.59 | 0.17 | A1 | 1.21 | 0.41 | 1 |

**Table S6. Meta-analysis of the replicated exonic associations.** Effect corresponds to OR (odds ratio) for logistic regression on AD status not adjusted by age (LogReg), exp(β) for linear regression on AD-age score (LinReg), and 1/HR (hazard ratio) for multivariate Cox regression on age-at-onset (CoxReg). In every model, σ corresponds to the standard error of the parameter estimate (i.e, log(OR) for LogReg, β for LinReg, and log(HR) for CoxReg).

**Part A.**

| SNP (hg19) / Gene | Discovery | | | | | | Replication | | | | | |
|---|---|---|---|---|---|---|---|---|---|---|---|---|
| | LogReg | | LinReg | | CoxReg | | LogReg | | LinReg | | CoxReg | |
| | **OR** | **σ** | **exp(β)** | **σ** | **1/HR** | **σ** | **OR** | **σ** | **exp(β)** | **σ** | **1/HR** | **σ** |
| 1:200959302:G:A / ***KIF21B*** | 0.87 | 0.04 | 0.90 | 0.02 | 0.89 | 0.02 | 0.96 | 0.03 | 0.96 | 0.02 | 0.96 | 0.02 |
| 1:216270469:G:A / ***USH2A*** | 9.12 | 0.76 | 6.76 | 0.35 | 4.07 | 0.24 | 1.58 | 0.31 | 1.70 | 0.27 | 1.33 | 0.19 |
| 2:26332640:T:C / ***RAB10*** | 17.43 | 1.52 | 10.46 | 0.34 | 4.92 | 0.32 | 4.50 | 0.76 | 5.03 | 0.48 | 2.69 | 0.29 |
| 6:41129252:C:T / ***TREM2*** | 4.83 | 0.25 | 3.22 | 0.10 | 2.58 | 0.09 | 2.32 | 0.14 | 2.69 | 0.12 | 1.95 | 0.08 |
| 7:99971313:T:C / ***PILRA*** | 0.88 | 0.03 | 0.87 | 0.03 | 0.90 | 0.02 | 0.92 | 0.02 | 0.90 | 0.02 | 0.93 | 0.01 |
| 11:59945745:T:C / ***MS4A6A*** | 0.88 | 0.03 | 0.91 | 0.02 | 0.92 | 0.02 | 0.89 | 0.02 | 0.89 | 0.02 | 0.93 | 0.01 |
| 14:93022240:G:T / ***RIN3*** | 16.32 | 1.04 | 6.54 | 0.32 | 3.46 | 0.24 | 1.95 | 0.33 | 1.69 | 0.23 | 1.59 | 0.18 |
| 16:29998200:A:G / ***TAOK2*** | 1.12 | 0.03 | 1.08 | 0.02 | 1.09 | 0.02 | 1.04 | 0.02 | 1.05 | 0.02 | 1.05 | 0.01 |
| 17:44828931:G:A / ***NSF/MAPT/KANSL1*** | 0.85 | 0.04 | 0.89 | 0.02 | 0.89 | 0.02 | 0.97 | 0.03 | 0.97 | 0.02 | 0.98 | 0.02 |
| 19:1047507:AGGAGCAG:A / ***ABCA7*** | 3.36 | 0.31 | 2.18 | 0.16 | 1.94 | 0.14 | 1.36 | 0.20 | 1.33 | 0.18 | 1.22 | 0.13 |

**Part B.**

| SNP (hg19) / Gene | Meta-analysis | | | | | | | | |
|---|---|---|---|---|---|---|---|---|---|
| | LogReg | | | LinReg | | | CoxReg | | |
| | **OR** | **σ** | **p** | **exp(β)** | **σ** | **p** | **1/HR** | **σ** | **p** |
| 1:200959302:G:A / ***KIF21B*** | 0.93 | 0.02 | $9.6.10^{-4}$ | 0.93 | 0.02 | $2.1.10^{-5}$ | 0.94 | 0.01 | $3.6.10^{-6}$ |
| 1:216270469:G:A / ***USH2A*** | 2.02 | 0.28 | $1.3.10^{-2}$ | 2.90 | 0.21 | $7.1.10^{-7}$ | 2.01 | 0.15 | $2.2.10^{-6}$ |
| 2:26332640:T:C / ***RAB10*** | 5.92 | 0.68 | $9.3.10^{-3}$ | 8.16 | 0.28 | $3.1.10^{-14}$ | 3.54 | 0.21 | $3.4.10^{-9}$ |
| 6:41129252:C:T / ***TREM2*** | 2.76 | 0.12 | $8.8.10^{-17}$ | 3.00 | 0.08 | $1.6.10^{-45}$ | 2.17 | 0.06 | $2.5.10^{-39}$ |
| 7:99971313:T:C / ***PILRA*** | 0.91 | 0.02 | $1.1.10^{-8}$ | 0.89 | 0.02 | $1.9.10^{-12}$ | 0.92 | 0.01 | $3.2.10^{-12}$ |
| 11:59945745:T:C / ***MS4A6A*** | 0.89 | 0.02 | $5.0.10^{-13}$ | 0.90 | 0.01 | $6.8.10^{-13}$ | 0.92 | 0.01 | $1.3.10^{-12}$ |
| 14:93022240:G:T / ***RIN3*** | 2.37 | 0.31 | $6.0.10^{-3}$ | 2.69 | 0.19 | $1.4.10^{-7}$ | 2.11 | 0.15 | $4.0.10^{-7}$ |
| 16:29998200:A:G / ***TAOK2*** | 1.07 | 0.02 | $9.8.10^{-5}$ | 1.06 | 0.01 | $1.3.10^{-6}$ | 1.06 | 0.01 | $2.9.10^{-8}$ |
| 17:44828931:G:A / ***NSF/MAPT/KANSL1*** | 0.92 | 0.02 | $1.9.10^{-4}$ | 0.93 | 0.02 | $7.8.10^{-6}$ | 0.95 | 0.01 | $6.2.10^{-5}$ |
| 19:1047507:AGGAGCAG:A / ***ABCA7*** | 1.77 | 0.17 | $6.5.10^{-4}$ | 1.75 | 0.12 | $3.2.10^{-6}$ | 1.52 | 0.09 | $7.9.10^{-6}$ |

**Table S7. Brain *cis*-eQTL associations with common synonymous variants reported in Table 2.** We queried the largest brain *cis*-eQTL meta-analysis which included 1,433 post-mortem brain samples from the AMP-AD and CommonMind Consortium. Only nominal significant eQTL associations ($p < 0.05$) and associations with mapped gene are reported below. Associations are sorted by locus and significance, in bold is the eQTL association with the mapped gene(s). (DOI: 10.7303/syn16984815.1).

| SNP / Mapped gene | Gene | P-val | FDR | Beta | Expression Increasing Allele | Gene Biotype |
|---|---|---|---|---|---|---|
| *rs2297911 / KIF21B* | *TMEM9* | 3.00E-02 | 3.36E-01 | -0.10 | G | protein coding |
| *rs2297911 / KIF21B* | *TIMM17A* | 4.08E-02 | 3.92E-01 | -0.09 | G | protein coding |
| *rs2297911 / KIF21B* | *DDX59* | 4.09E-02 | 3.92E-01 | -0.10 | G | protein coding |
| ***rs2297911 / KIF21B*** | ***KIF21B*** | **4.71E-02** | **4.20E-01** | **0.09** | **A** | **protein coding** |
| *rs2405442 / PILRA* | *PVRIG* | 1.05E-24 | 2.00E-22 | -0.52 | T | protein coding |
| *rs2405442 / PILRA* | *STAG3L5P* | 3.83E-14 | 3.87E-12 | 0.31 | C | transcribed unprocessed pseudogene |
| *rs2405442 / PILRA* | *PILRB* | 2.12E-13 | 2.03E-11 | 0.31 | C | protein coding |
| *rs2405442 / PILRA* | *STAG3* | 1.71E-11 | 1.41E-09 | -0.26 | T | protein coding |
| ***rs2405442 / PILRA*** | ***PILRA*** | **8.61E-07** | **3.92E-05** | **0.21** | **C** | **protein coding** |
| *rs2405442 / PILRA* | *AP4M1* | 2.95E-06 | 1.22E-04 | -0.19 | T | protein coding |
| *rs2405442 / PILRA* | *TRIM4* | 4.29E-06 | 1.71E-04 | -0.19 | T | protein coding |
| *rs2405442 / PILRA* | *MBLAC1* | 3.46E-05 | 1.14E-03 | 0.17 | C | protein coding |
| *rs2405442 / PILRA* | *GIGYF1* | 8.43E-05 | 2.54E-03 | -0.16 | T | protein coding |
| *rs2405442 / PILRA* | *AZGP1* | 4.77E-04 | 1.17E-02 | -0.15 | T | protein coding |
| *rs2405442 / PILRA* | *PMS2P1* | 7.85E-03 | 1.14E-01 | -0.11 | T | unprocessed pseudogene |
| *rs2405442 / PILRA* | *GATS* | 1.75E-02 | 1.98E-01 | -0.10 | T | protein coding |
| *rs2405442 / PILRA* | *MCM7* | 1.75E-02 | 1.98E-01 | -0.10 | T | protein coding |
| *rs2405442 / PILRA* | *MEPCE* | 2.51E-02 | 2.49E-01 | -0.09 | T | protein coding |

| | | | | | | |
|---|---|---|---|---|---|---|
| *rs2405442 / PILRA* | *VGF* | 2.72E-02 | 2.61E-01 | 0.09 | C | protein coding |
| *rs2405442 / PILRA* | *SLC12A9* | 3.49E-02 | 3.03E-01 | -0.09 | T | protein coding |
| *rs2405442 / PILRA* | *TSC22D4* | 3.66E-02 | 3.11E-01 | -0.09 | T | protein coding |
| *rs2405442 / PILRA* | *AP1S1* | 4.09E-02 | 3.31E-01 | 0.09 | C | protein coding |
| *rs2405442 / PILRA* | *MUC12* | 4.58E-02 | 3.52E-01 | 0.09 | C | protein coding |
| *rs12453 / MS4A6A* | *PRPF19* | 2.85E-03 | 7.27E-02 | -0.11 | T | protein coding |
| *rs12453 / MS4A6A* | *MS4A4A* | 6.12E-02 | 4.71E-01 | -0.10 | T | protein coding |
| ***rs12453 / MS4A6A*** | ***MS4A6A*** | **5.17E-01** | **8.98E-01** | **-0.02** | **T** | **protein coding** |
| *rs4077410 / TAOK2* | *INO80E* | 3.10E-80 | 5.95E-77 | -0.62 | A | protein coding |
| *rs4077410 / TAOK2* | *SMG1P5* | 1.27E-11 | 1.60E-09 | 0.32 | G | transcribed unprocessed pseudogene |
| *rs4077410 / TAOK2* | *MAPK3* | 1.40E-09 | 1.40E-07 | -0.22 | A | protein coding |
| *rs4077410 / TAOK2* | *TBX6* | 1.43E-09 | 1.43E-07 | 0.29 | G | protein coding |
| *rs4077410 / TAOK2* | *TMEM219* | 7.67E-08 | 6.01E-06 | -0.20 | A | protein coding |
| *rs4077410 / TAOK2* | *PPP4C* | 6.26E-06 | 3.54E-04 | -0.17 | A | protein coding |
| *rs4077410 / TAOK2* | *ENSG00000250616* | 7.37E-06 | 4.10E-04 | -0.17 | A | antisense |
| *rs4077410 / TAOK2* | *YPEL3* | 1.10E-05 | 5.88E-04 | 0.16 | G | protein coding |
| *rs4077410 / TAOK2* | *NPIPB12* | 3.18E-04 | 1.17E-02 | 0.13 | G | protein coding |
| *rs4077410 / TAOK2* | *GDPD3* | 3.69E-03 | 8.80E-02 | -0.11 | A | protein coding |
| *rs4077410 / TAOK2* | *KCTD13* | 9.57E-03 | 1.73E-01 | -0.09 | A | protein coding |
| *rs4077410 / TAOK2* | *CDIPT* | 1.38E-02 | 2.19E-01 | 0.09 | G | protein coding |
| *rs4077410 / TAOK2* | *DOC2A* | 1.56E-02 | 2.36E-01 | -0.09 | A | protein coding |
| *rs4077410 / TAOK2* | *ASPHD1* | 1.67E-02 | 2.46E-01 | 0.09 | G | protein coding |
| ***rs4077410 / TAOK2*** | ***TAOK2*** | **2.67E-02** | **3.24E-01** | **-0.08** | **A** | **protein coding** |
| *rs4077410 / TAOK2* | *ITGAL* | 2.95E-02 | 3.42E-01 | -0.08 | A | protein coding |

| | | | | | | |
|---|---|---|---|---|---|---|
| *rs4077410 / TAOK2* | *CORO1A* | 3.18E-02 | 3.56E-01 | -0.08 | A | protein coding |
| *rs4077410 / TAOK2* | *C16orf92* | 3.32E-02 | 3.64E-01 | -0.19 | A | protein coding |
| *rs4077410 / TAOK2* | *TBC1D10B* | 3.49E-02 | 3.73E-01 | 0.08 | G | protein coding |
| *rs4077410 / TAOK2* | *NPIPB11* | 4.10E-02 | 4.04E-01 | -0.07 | A | protein coding |
| *rs199533 / NSF/MAPT/KANSL1* | *KANSL1-AS1* | 8.99E-214 | 1.60E-210 | 1.45 | A | antisense |
| *rs199533 / NSF/MAPT/KANSL1* | *ARL17A* | 5.47E-185 | 3.87E-182 | 1.12 | A | protein coding |
| *rs199533 / NSF/MAPT/KANSL1* | *LRRC37A* | 1.56E-145 | 5.67E-143 | 1.05 | A | protein coding |
| *rs199533 / NSF/MAPT/KANSL1* | *LRRC37A2* | 1.64E-136 | 5.89E-134 | 1.01 | A | protein coding |
| ***rs199533 / NSF/MAPT/KANSL1*** | ***KANSL1*** | **6.65E-84** | **1.65E-81** | **0.85** | **A** | **protein coding** |
| *rs199533 / NSF/MAPT/KANSL1* | *ENSG00000262879* | 4.27E-24 | 5.47E-22 | 0.48 | A | processed transcript |
| *rs199533 / NSF/MAPT/KANSL1* | *CRHR1* | 1.70E-18 | 1.74E-16 | 0.55 | A | protein coding |
| *rs199533 / NSF/MAPT/KANSL1* | *WNT3* | 3.18E-13 | 2.59E-11 | 0.36 | A | protein coding |
| *rs199533 / NSF/MAPT/KANSL1* | *STH* | 2.29E-10 | 1.48E-08 | 0.32 | A | protein coding |
| *rs199533 / NSF/MAPT/KANSL1* | *SPPL2C* | 1.89E-09 | 1.09E-07 | 0.76 | A | protein coding |
| *rs199533 / NSF/MAPT/KANSL1* | *MAPT-IT1* | 2.84E-09 | 1.62E-07 | 0.38 | A | sense intronic |
| *rs199533 / NSF/MAPT/KANSL1* | *ENSG00000262881* | 2.73E-04 | 7.81E-03 | -0.19 | G | antisense |
| *rs199533 / NSF/MAPT/KANSL1* | *LRRC37A17P* | 1.16E-03 | 2.77E-02 | -0.16 | G | transcribed unprocessed pseudogene |
| *rs199533 / NSF/MAPT/KANSL1* | *ARL17B* | 1.02E-02 | 1.53E-01 | 0.12 | A | protein coding |
| *rs199533 / NSF/MAPT/KANSL1* | *ENSG00000274883* | 3.59E-02 | 3.38E-01 | 0.15 | A | misc RNA |
| ***rs199534 / NSF/MAPT/KANSL2*** | ***MAPT*** | **2.58E-01** | **7.62E-01** | **0.06** | **A** | **protein coding** |
| ***rs199535 / NSF/MAPT/KANSL3*** | ***NSF*** | **3.31E-01** | **8.12E-01** | **0.05** | **A** | **protein coding** |

**Table S7. Differential expression between AD and control individuals for mapped genes reported in Table 2.** We queried the AMP-AD fixed-effect meta-analysis of differential expression between AD and control individuals across brain tissues in the ROSMAP, MAYO and MSBB databases (DOI: 10.7303/syn11914606).

| Gene | TE | σ | lowerTE | upperTE | Z | P-val | Q | tau | H | I2 | FDR |
|---|---|---|---|---|---|---|---|---|---|---|---|
| *TREM2* | 0.37 | 0.07 | 0.24 | 0.50 | 5.67 | 1.43E-08 | 6.37 | 0.02 | 1.03 | 0.06 | 3.71E-07 |
| *TAOK2* | -0.37 | 0.07 | -0.49 | -0.24 | -5.55 | 2.83E-08 | 8.14 | 0.10 | 1.16 | 0.26 | 6.41E-07 |
| *KANSL1* | 0.32 | 0.07 | 0.20 | 0.45 | 4.93 | 8.23E-07 | 19.95 | 0.26 | 1.82 | 0.70 | 1.02E-05 |
| *RAB10* | 0.32 | 0.07 | 0.19 | 0.44 | 4.82 | 1.47E-06 | 5.56 | 0.04 | 1.00 | 0.00 | 1.64E-05 |
| *MS4A6A* | 0.22 | 0.07 | 0.09 | 0.35 | 3.40 | 6.79E-04 | 6.96 | 0.09 | 1.08 | 0.14 | 2.64E-03 |
| *RIN3* | 0.18 | 0.07 | 0.05 | 0.30 | 2.67 | 7.48E-03 | 15.52 | 0.22 | 1.61 | 0.61 | 1.95E-02 |
| *PILRA* | 0.14 | 0.07 | 0.01 | 0.27 | 2.13 | 3.30E-02 | 15.26 | 0.22 | 1.59 | 0.61 | 6.70E-02 |
| *ABCA7* | 0.11 | 0.07 | -0.02 | 0.24 | 1.70 | 8.87E-02 | 7.67 | 0.10 | 1.13 | 0.22 | 1.51E-01 |
| *MAPT* | -0.11 | 0.07 | -0.24 | 0.02 | -1.65 | 9.89E-02 | 10.06 | 0.14 | 1.30 | 0.40 | 1.66E-01 |
| *NSF* | -0.10 | 0.07 | -0.23 | 0.03 | -1.53 | 1.27E-01 | 16.70 | 0.23 | 1.67 | 0.64 | 2.03E-01 |
| *KIF21B* | -0.09 | 0.07 | -0.21 | 0.04 | -1.30 | 1.92E-01 | 19.28 | 0.25 | 1.79 | 0.69 | 2.83E-01 |
